# Social Determinants of Health Factors Associated with Metabolic Dysfunction-Associated Steatotic Liver Disease Prevalence and Severity: A Systematic Review and Meta-analysis

**DOI:** 10.1101/2024.09.29.24314567

**Authors:** Mohammed Abu-Rumaileh, Sudheer Dhoop, Jordan Pace, Thabet Qapaja, Maria Elena Martinez, Monica Tincopa, Rohit Loomba

## Abstract

**Background & Aims:** Social determinants of health (SDOH) impact disease risk and severity leading to health disparities and impeding health equity. Though important in mitigating adverse health outcomes, SDOH impacting metabolic dysfunction-associated steatotic liver disease (MASLD) prevalence and severity are understudied and results are conflicting. The aim of this systematic review and meta-analysis was to assess the impact of specific SDOH factors on MASLD disease burden for adults in the United States (US).

**Methods:** We searched MEDLINE, Embase and Cochrane databases per the Preferred Reporting Items for Systematic Reviews and Meta-analyses guidelines. Studies from January 2010-May 2024 were included. Quantitative studies of adults in the US that evaluated SDOH beyond race/ethnicity were included. Outcomes included prevalence of MASLD, metabolic dysfunction-associated steatohepatitis (MASH), MASH-associated advanced fibrosis or cirrhosis and clinical outcomes.

**Results:** We identified 18 studies comprising of 547,634 total subjects from 11 unique cohorts. Nine studies evaluated MASLD prevalence, three MASH prevalence, eight MASH-associated advanced fibrosis/cirrhosis prevalence, and nine clinical outcomes. High diet quality was the most consistent SDOH factor associated with both MASLD and MASH-associated advanced fibrosis/cirrhosis prevalence (summarized OR of 0.76 p <0.01, and 0.74 p <0.01, respectively). Lower income was most consistently associated with risk of clinical outcomes (significant in 3/9 studies).

**Conclusions:** Diet quality was the most consistent SDOH associated with disease prevalence and severity in MASLD, with the remainder of SDOH showing inconsistent associations. Prospective assessments using consistent, validated tools to assess the impact of specific SDOH on MASLD disease burden are needed to inform public health interventions to mitigate health disparities in MASLD.

## INTRODUCTION

Metabolic dysfunction-associated steatotic liver disease (MASLD) is characterized by hepatic fat accumulation without secondary causes such as significant alcohol consumption, drug-induced steatosis, or inherited disorders. [1] [2] It can progress histologically from simple steatosis to steatohepatitis [metabolic dysfunction-associated steatohepatitis (MASH)], advanced fibrosis and cirrhosis, with risk for development of hepatocellular carcinoma (HCC) and need for liver transplantation. [3] MASLD is a highly prevalent disease that is estimated to affect over 30% of adults the United States (US) and the worldwide population. [4, 5] MASH is projected to be the leading cause of decompensated liver disease and liver transplantation based on trends in liver transplantation in the U.S. [6-9] Given this trend, it is imperative to identify underlying mediators for the development and progression of MASLD.

Numerous studies have characterized genetic factors associated with MASLD disease prevalence and severity. The patatin-like phospholipase domain-containing protein 3 (PNPLA3) polymorphism is among one of the key genetic factors identified in mediating MASLD/MASH disease burden and severity. [10-12] In addition to genetic factors, dietary and exercise patterns have also been extensively evaluated with respect to risk of MASLD and MASH. Higher intake of total calories, red and processed meats, high fructose corn syrup and carbohydrates have been associated with increased risk of MASLD and MASH. [13] Lower rates of physical activity have also been correlated with risk of MASLD and MASH. [14, 15] Detailed analyses of other environmental, structural and social factors that impact MASLD disease development have been limited, though the body of research on this topic is increasing. The concept of social determinants of health (SDOH) provides a useful framework for understanding the social and environmental factors influencing the prevalence and outcomes of chronic diseases. [16-19] SDOH are defined as conditions people are born into or encounter throughout their lives. SDOH domains include, but are not limited to, economic stability, education access and quality, health care access and quality, neighborhood and built environment, and social and community context. [20] Adverse SDOH disproportionately impact underserved and minority populations, leading to health disparities with respect to higher rates of disease and adverse health outcomes. [21] Underlying SDOH are hypothesized as being causative drivers in health disparities seen in MASLD and MASH with minority populations including Hispanic/Latino individuals experiencing higher MASLD and MASH disease prevalence. [22] Therefore, it is critical to more definitively characterize the relationship between SDOH and MASLD disease prevalence and severity in order to inform public health efforts in MASLD to mitigate health disparities and improve overall health outcomes for vulnerable populations with a goal of achieving health equity.[23-25]

Though crucially important to evaluate to mitigate adverse health outcomes, detailed analyses of SDOH impacting MASLD prevalence, severity and clinical outcomes are limited, and results are contradictory. [26] The majority of the existing literature evaluating SDOH in MASLD are retrospective database analyses or analyses of the same database [National Health and Nutrition Examination Survey (NHANES)]. [27] Across the existing literature, there is a lack of uniformity in definition of MASLD and MASH. There is also heterogeneity with respect to the SDOH variables studied and definitions of these variables across studies. Few studies systematically evaluate well defined SDOH variables beyond race/ethnicity. These characteristics pose difficulties in ascertaining a clear understanding of which factors would be highest yield to target for directed public health and policy interventions. Therefore, the aim of this systematic review and meta-analysis is to identify the association between individual SDOH factors beyond race and ethnicity and MASLD disease prevalence, severity and clinical outcomes for adults in the US.

## METHODS

### Data Sources and Search Strategy

We followed the Preferred Reporting Items for Systematic reviews and Meta-Analyses (PRISMA) recommendations in conducting this systematic review and meta-analysis. [28] With the assistance of a medical research librarian, we performed serial literature searches for articles of interest. MEDLINE (via PubMed), EMBASE, and Cochran were searched using the following keywords: “Non-alcoholic Fatty Liver Disease”, “fatty liver disease”, “Steatohepatitis”, “Steatonecrosis”, “Nonalcoholic steatohepatitis”, “metabolic dysfunction-associated steatotic liver disease”, “MASLD”, “NAFLD”, “social determinants of health”, “Socioeconomic disparities”, “Underserved populations”, “Social determinants of health”, “Income”, “food insecurity”, “housing”, “Universal coverage”, “Universal healthcare”, “Socioeconomic status”, “Health Inequity” “urban health”, “vulnerable population”, “social deprivation index”, “disparities”, “Racial disparities”, “Distance”, “urban”, “Universal health coverage”, “Socioeconomic position”, “Inequities”, “Equity”, “Inequalities”, and “healthcare access”. Boolean operators and medical subject heading (MESH) terms as well as other controlled vocabulary were used to enhance electronic searches. The specific search strategy is shown in **Supplemental File 1**. Additional studies of interest were identified by cited reference tracking, hand searched of bibliographies, and consultation with clinical experts on the topic. The initial search was performed in November 2023 and was updated in May 2024.

### Study Eligibility and Selection Criteria

Two study authors determined study eligibility. Studies were initially screened by the first author. Decisions about study inclusion were made independently by two authors (M.A. and S.D.). Differences in opinion regarding study inclusion were resolved through consensus. Final study inclusion eligibility was confirmed by the senior author (M.T.). Original, adult (age 18 or older) human patient studies from the US that systematically evaluated the impact of SDOH on prevalence of MASLD, MASH, MASH-associated advanced fibrosis/cirrhosis, and clinical outcomes in MASLD/MASH were eligible for inclusion. For outcome definition of MASLD or MASH, studies used liver biopsy, imaging, combination imaging and serologic parameters and international classification of diseases (ICD)-9 codes. We excluded studies that were not done on the US population, done in the pediatric population or animals, done only within specific subpopulations with MASLD (i.e. sex, race or ethnicity), included patients with other forms of liver disease, done in overlapping cohorts, and studies that evaluated race/ethnicity as the only SDOH factor. Study published in 2010 and beyond were eligible for inclusion as this most accurately reflects the current disease prevalence for MASLD and MASH in the setting of the evolving obesity epidemic. Lastly, studies for which no translation into English language was available were also excluded.

### Data Abstraction and Validity Assessment

Data from eligible studies were abstracted by 2 authors independently to ensure data abstraction accuracy. For all studies, we recorded the study design, sample size, patient population characteristics, SDOH variables studied, outcomes measures, criteria to define outcomes, and measures of association/prediction of risk for outcomes. A variable was classified as a SDOH based on the NIH framework and inclusion in the National Institute on Minority Health’s (NIMHD) recommended assessment strategy for SDOH, PhenX.[29] We accepted the outcome definition as stated by each study without independently validating or reviewing their data. Study authors were directly contacted for additional information when necessary. Study quality was assessed using the NIH Quality Assessment Tool for Observational Cohort and Cross-sectional Studies independently by two authors. [30]

### Data Synthesis and Analysis

Two authors synthesized the results of the included studies. Studies were categorized according to the outcome of interest (MASLD, MASH, MASH-associated advanced fibrosis/cirrhosis, or clinical outcomes). For SDOH factors that were evaluated in multiple studies and noted to have significant association with outcomes of interest, meta-analysis was performed. Meta-analysis was performed using a random effects model to assess summarized odds ratios (ORs) based on results of multivariate analysis of predictors. SDOH factors were selected for meta-analysis based on the number of studies these factors were evaluated in and the amenability of the data to be included in a meta-analysis (i.e. factor defined similarly across studies). Publication bias was assessed using funnel plots. P <0.05 was considered statistically significant. Heterogeneity was assessed using the I2 statistic with a cutoff value of >75% representing substantial heterogeneity, as per the Cochrane handbook.

## RESULTS

### Studies Included in the Systematic Review

Our search method identified 4764 studies, of which 3845 were screened after removal of duplicated and studies done on animals (**Figure 1**). Two authors independently assessed 3210 unique articles for eligibility based on title and abstract. Of these, 2846 were excluded due to study location outside of the US (n=574), focused on management/intervention (n=414), included alternative etiologies of liver disease (n=187), or was not related to SDOH in MASLD (n=1671). Of the remaining 364 studies, 18 articles met the predefined criteria for analysis.

**Figure 1.**
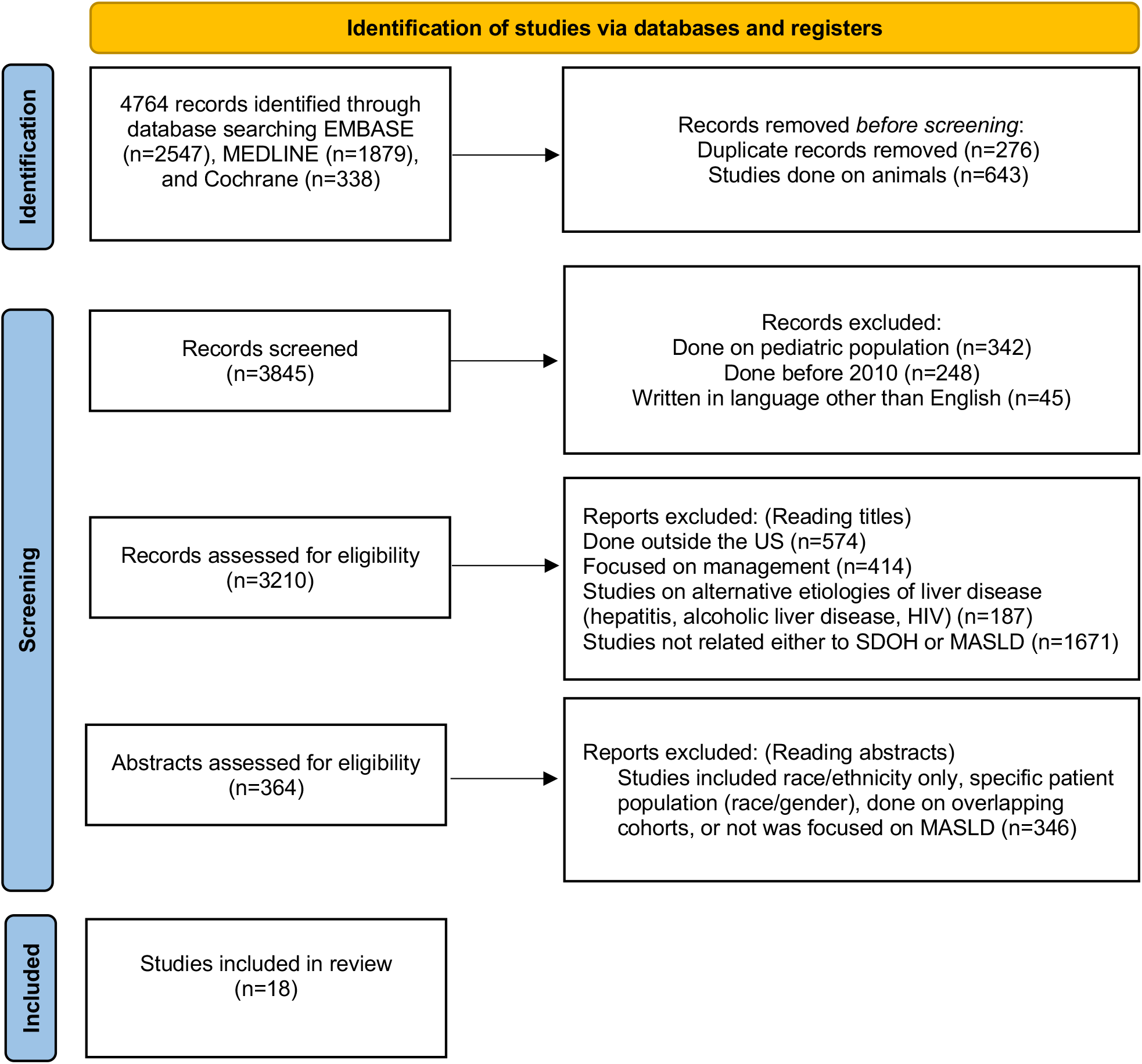
PRISMA 2020 flow diagram for systematic review.

**Figure 2.**
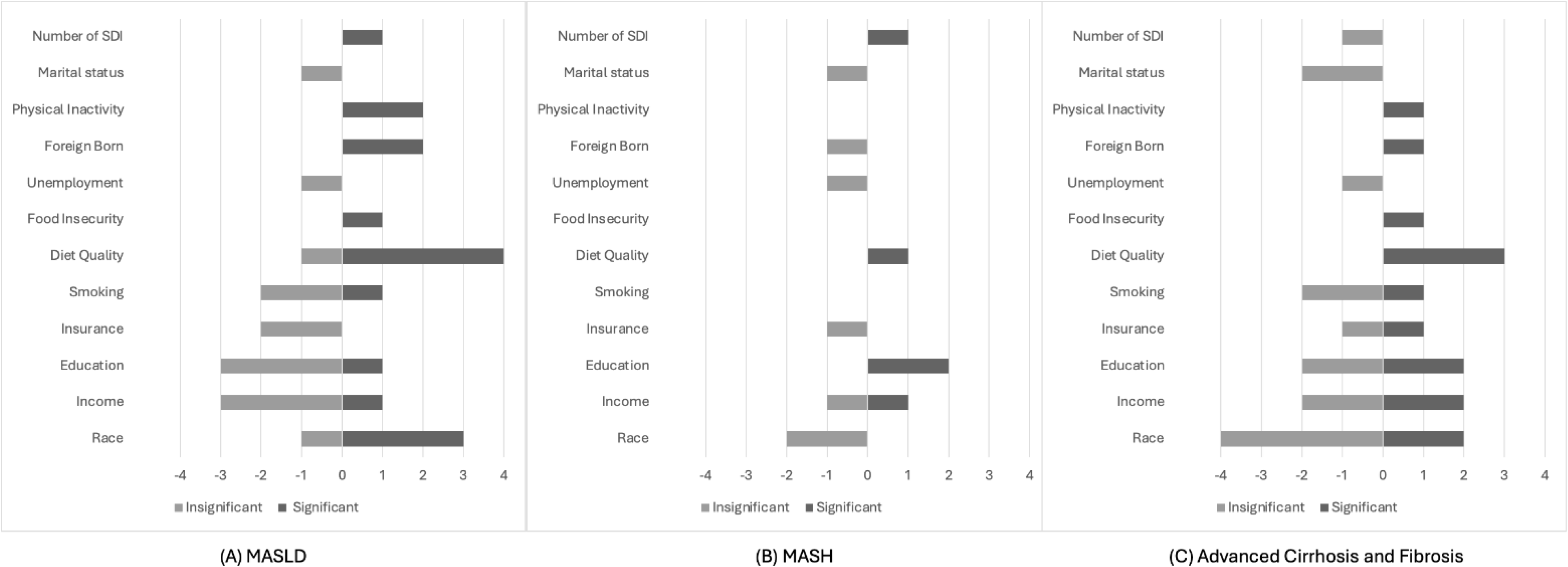
Y-axis shows variables identified to be predictive of MASLD, MASH, and advanced fibrosis and cirrhosis prevalence. X-xis shows the number of studie reporting that outcome.

A total of 547,634 total subjects from 11 unique patient cohorts were represented. Eight of the studies used the NHANES cohort. Nine studies evaluated MASLD prevalence, three studies evaluated MASH prevalence, eight studies evaluated MASH-associated advanced fibrosis/cirrhosis prevalence, and nine studies evaluated clinical outcomes. All studies included univariate or multivariate analyses. MASLD prevalence was diagnosed based on liver biopsy (n=1), US fatty liver index (USFLI) (n=3), fibroscan (n=3), ICD codes (n=1), and ultrasound imaging (n=1) as shown in **Table 1**. MASH prevalence was defined based on liver biopsy (n=2) and fibroscan (n=1) as shown in **Table 2**. Advanced fibrosis and cirrhosis prevalence were defined by liver biopsy (n=2), NAFLD fibrosis score (NFS) (n=2), fibroscan (n=2), magnetic resonance elastography (MRE) (n=1), and ICD codes (n=1) as shown in **Table 3**. Clinical outcomes evaluated were highly variable. The most commonly included clinical outcomes included overall mortality (n=5) and cardiovascular disease/events (n=3) (**Supplement Table 1**). The most common SDOH factors evaluated included race (n=9), income (n=9), education (n=7), insurance (n=5), smoking (n=5), diet quality (n=5), food insecurity (n=2), occupational status (n=2), foreign born (n=2), marital status (n=2), and physical activity (n=2) (**Supplement Table 2**).

### Quality Assessment and Risk of Bias

The quality of each individual study was assessed using the NIH Quality Assessment Tool for Observational Cohort and Cross-sectional Studies (**Supplement Table 3**). Eight studies were assessed as good quality with a score of >7, nine studies as fair quality with a score of 5-7, and one study as poor quality with a score of <5 . Of note, the study assessed as being poor quality was an abstract. Due to the nature of cross-sectional studies, the majority of studies were unable the satisfy the criteria for exposures of interest were measured prior to the outcomes being measured, timeframe was sufficient to reasonably expect to see an association between exposure and outcome if it existed, the exposures were assessed more than once overtime, and loss to follow-up was not reported. Funnel plot of studies with multivariate analysis did not note significant publication bias for studies amenable to meta-analysis (**Supplement Figure 1**).

### SDOH Factors Associated with MASLD Prevalence

A detailed list of SODH factors evaluated for association with MASLD disease prevalence is shown in **Table 1**. The most common SDOH factors found to be significantly associated MASLD prevalence were diet quality (n=4) [31-34], race (n=3) [35-37], physical inactivity (n=2) [37, 38], and foreign born status (n=2) [35, 39](significant in 4/5, 3/4, 2/2 and 2/2 studies respectively). Employment status, insurance type, and marital status were not found to be predictive of MASLD prevalence as shown in **Figure 2a**. Meta-analysis was performed to assess the impact of high diet quality (**Figure 3A**) and less than high school education (**Figure 3B**) on MASLD prevalence. High diet quality had a summarized OR of 0.76 (95% CI 0.67-0.86, p <0.01, I2 63%) and less than high school education had a summarized OR of 1.02 (95%CI 0.86-1.23, p=0.80, I2 62%).

**Figure 3.**
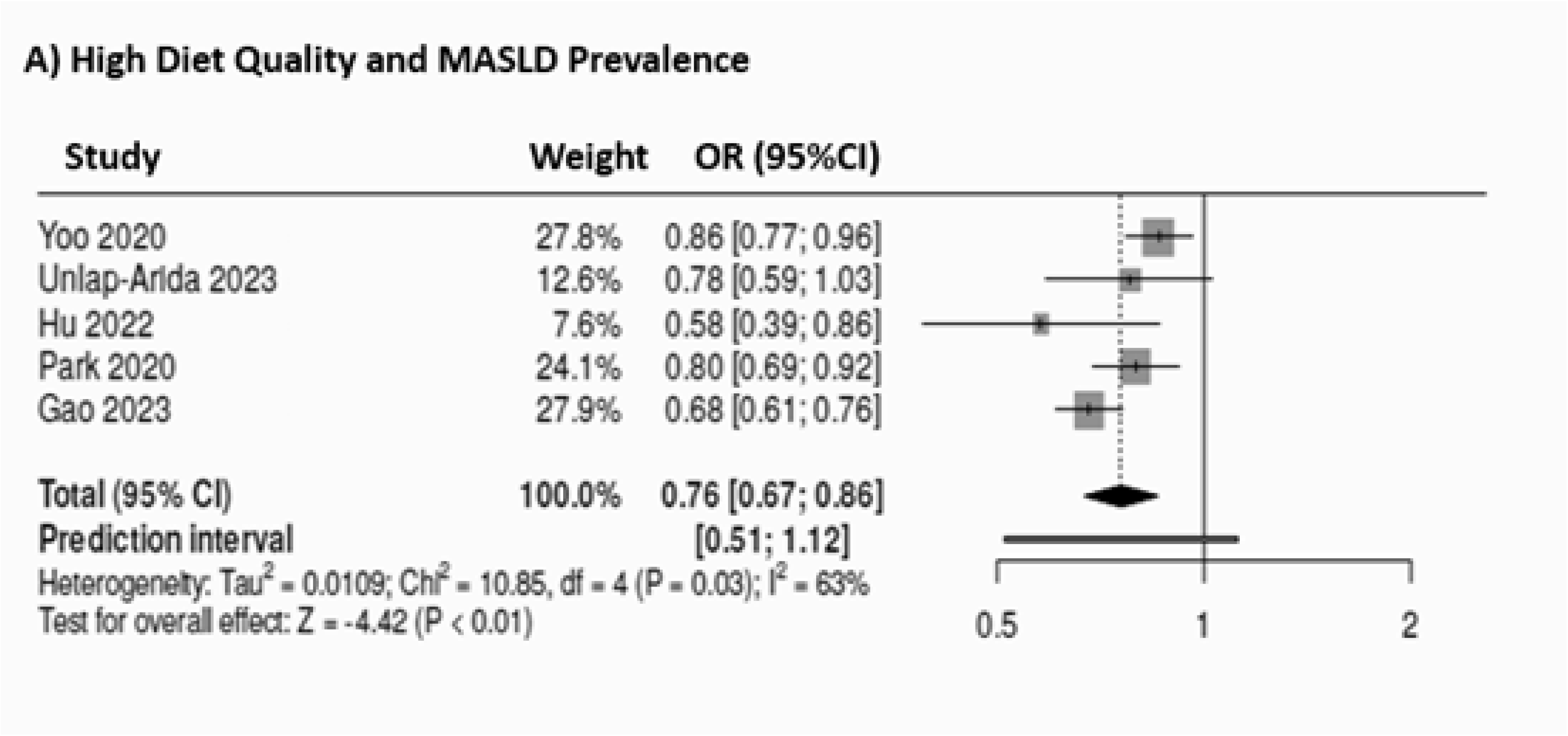
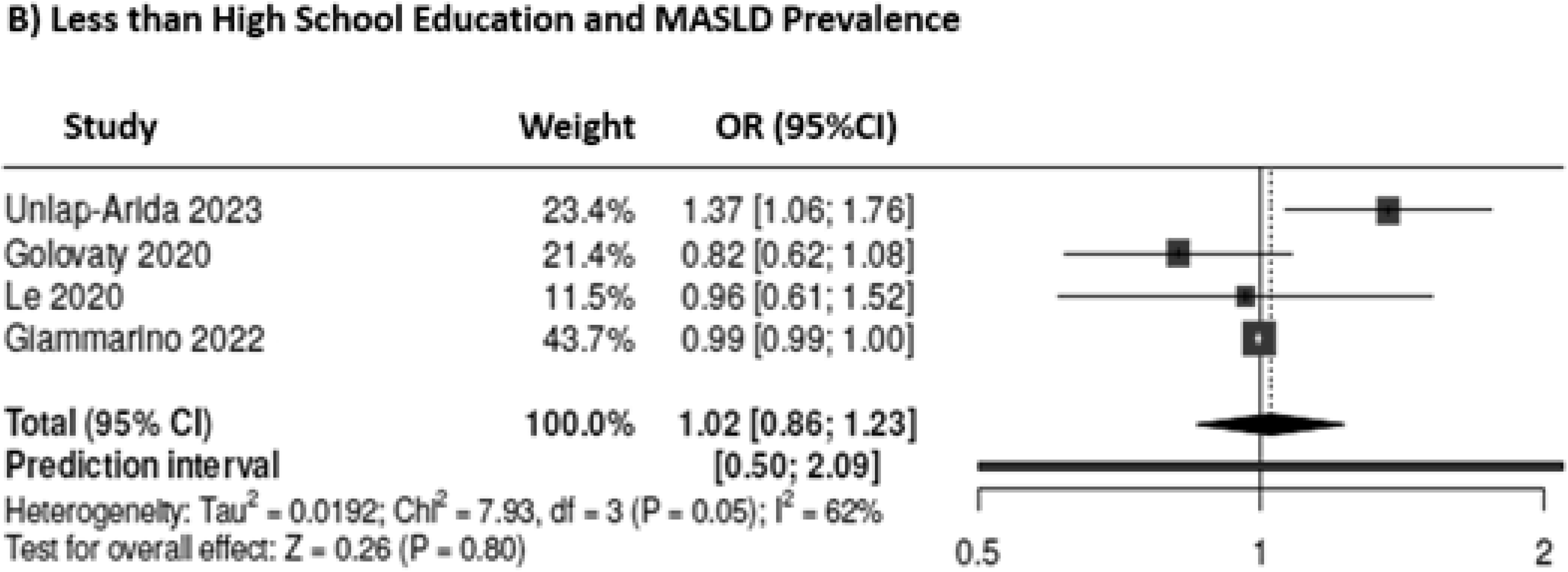
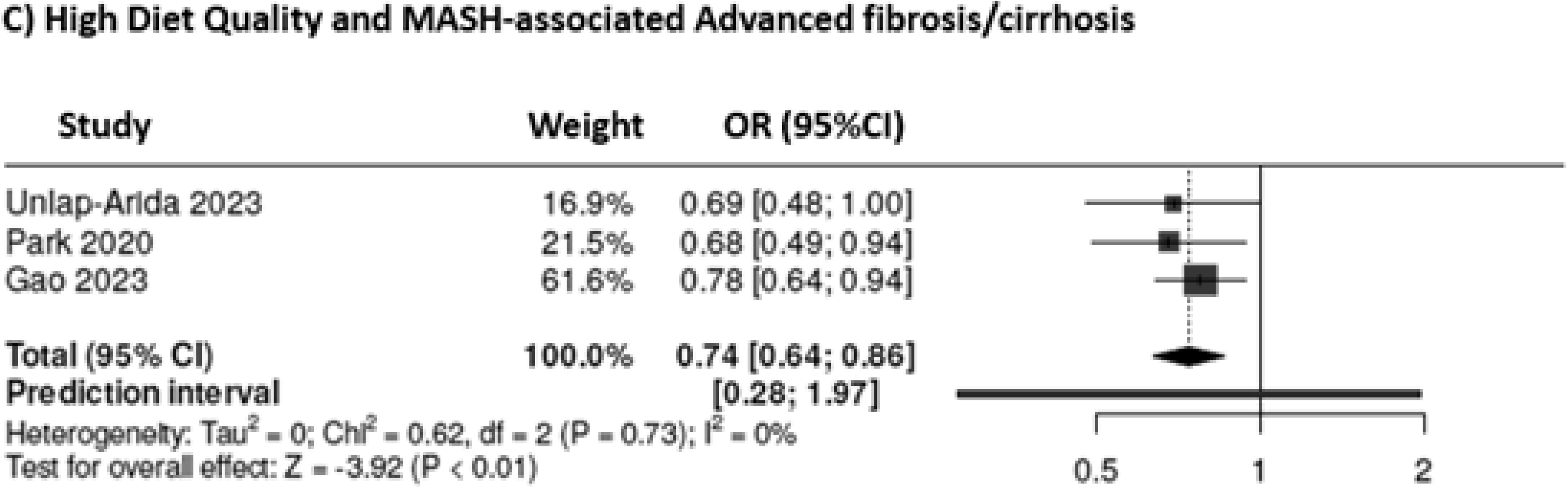
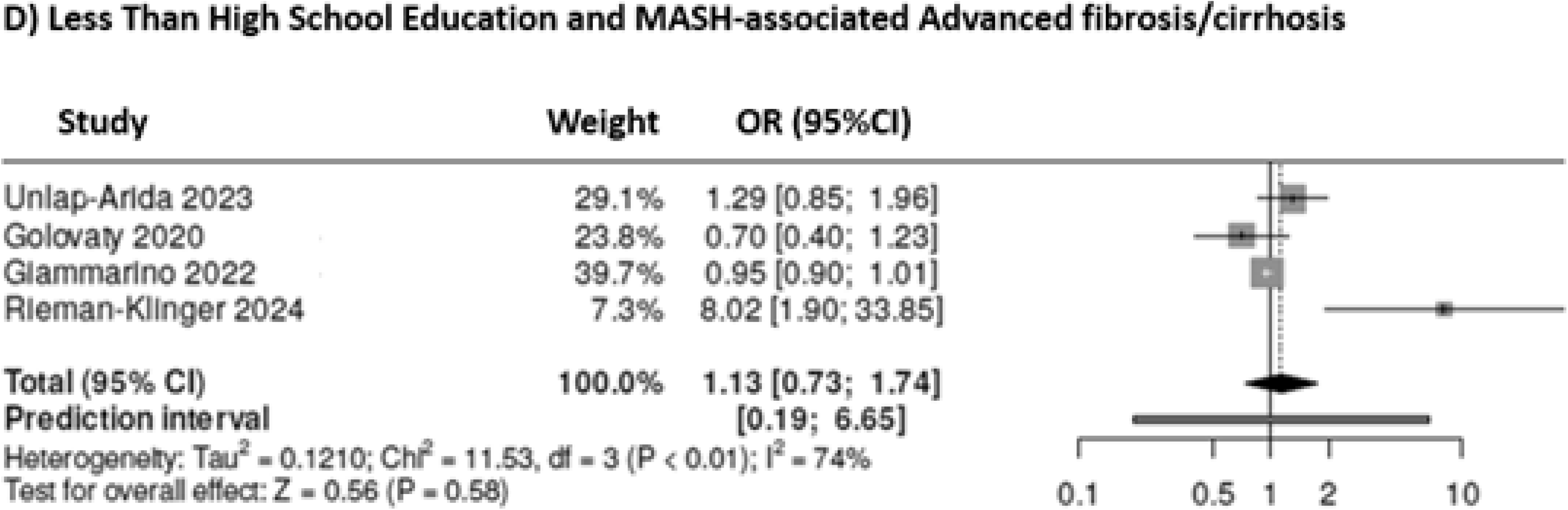
Forest Plot for Impact of. A) High Diet Quality and MASLD Prevalence B) Less than High School Education and MASLD Prevalence C)) High Diet Quality and MASH-associated Advanced fibrosis/cirrhosis and D) Less than High School Education and MASH-associated Advanced fibrosis/cirrhosis

### SDOH Factors Associated with MASH Prevalence

A detailed list of SODH factors evaluated for association with MASH disease prevalence is shown in **Table 2**. The most common SDOH factors found to be significantly associated with MASH prevalence were education (n=2) [39, 40], diet quality (n=1) [39], income (n=1) [40], and social deprivation index (SDI) score of +4 [39] (significant in 2/2, 1/1, 1/2, and 1/1 studies respectively). Race, insurance status, employment status, foreign born status, and marital status were not predictive of MASH prevalence as shown in **Figure 2b**. Given that no individual SDOH factor was found to be significant in 3 or more studies for this outcome of interest, meta-analysis was not possible.

### SDOH Factors Associated with MASH-Associated Advanced Fibrosis/Cirrhosis

A detailed list of SODH factors evaluated for association with MASH-associated advanced fibrosis and cirrhosis is shown in **Table 3**. The most common SDOH factors found to be significantly associated with MASH-associated advanced fibrosis/cirrhosis were diet quality (n=3) [31, 33, 37], race (n=2) [35, 36], income (n=2) [37, 41], and education (n=2) [37, 41] (significant in 3/3, 2/6, 2/4, and 2/4 studies respectively). Employment status, marital status, and SDI score were not significantly associated with MASH-associated advanced fibrosis/cirrhosis prevalence as shown in **Figure 1C**. Meta-analysis was performed to assess the impact of high diet quality (**Figure 3C**) and less than high school education (**Figure 3D**) on MASH-associated advanced fibrosis/cirrhosis. High diet quality had a summarized OR of 0.74 (95% CI 0.64-0.86, p <0.01, I2 0%) and less than high school education had a summarized OR of 1.13 (95%CI 0.73-1.74, p=0.58, I2 74%).

### SDOH Factors Associated with Clinical Outcomes in MASLD/MASH

A detailed list of SODH factors evaluated for association with clinical outcomes in MASLD/MASH is shown in **Supplement Table 1 and 2**. The clinical outcomes evaluated most often were overall mortality (n=5) and cardiovascular disease/events (n=3). Collectively, risk of overall mortality as well as risk of cardiovascular disease/events were most consistently associated with lower income (n=3 and n=2), with the remaining SDOH factors showing significant association with overall mortality in only single studies.

Higher income was associated with lower risk for both all-cause and cardiovascular related mortality [42-44], while low income was predictive for receiving healthcare 1-3 times per year [35] and increased length of inpatient stay [45]. Compared to private insurance, both publicly insured and uninsured individuals had higher all-cause mortality, unfavorable discharge disposition and increased length of stay. [42, 45]. Compared to full employment, part-time, retired, and unemployment were associated with fibrosis progression. [46] Foreign born status was associated with receiving less healthcare services compared to US born status. [35] Education above high school was associated with decreased risk for all-cause mortality.[42] Diet, exercise and other lifestyle behaviors also demonstrated significant associations with clinical outcomes. Food insecure patients were shown to have a higher risk for all-cause mortality and utilization of outpatient healthcare services. [42] Higher diet quality was associated with lower risk for all-cause and cancer-related mortality. [34] Physical activity was associated with decreased risk of mortality. [38] Smoking was found to be predictive for all-cause mortality and fibrosis progression. [42, 46]

## DISCUSSION

### Main Findings

MASLD prevalence is rapidly increasing and is a major cause of liver related morbidity and mortality both within the US and worldwide. While numerous potential risk factors have been associated with MASLD development and progression, detailed analysis of the influence of individual SDOH factors on MASLD disease burden beyond race/ethnicity remains limited and results have been conflicting. Our systematic review and meta-analysis addresses this knowledge gap by providing summative analysis on the impact of specific SDOH factors on MASLD, MASH and MASH-associated advanced fibrosis/cirrhosis prevalence and also highlights associations with clinical outcomes in MASLD. In this analysis we found that among variables assessed in multiple studies for risk of MASLD, diet quality, physical inactivity and foreign born status were most consistently associated with MASLD prevalence, with high diet quality having a summarized OR of 0.76 (95% CI 0.67-0.86, p <0.01). For SDOH associations and risk of MASH, education, income, diet, and SDI score were most consistent predictors. Risk of advanced fibrosis/cirrhosis was most consistently associated with diet quality, education, and income with high diet quality having a summarized OR of 0.74 (95% CI 0.64-0.86, p <0.01). SDOH factors associated with clinical outcomes were more challenging to assess given significant heterogeneity in outcomes assessed and definitions of outcomes, but lower income was most consistently associated with adverse clinical outcomes.

### In context with current literature

Our study builds on the existing literature by evaluating the impact of individual SDOH factors in MASLD across cohorts, accounting for conflicting results between studies. Overall it is well established that lifestyle behaviors such as diet quality and physical activity are associated with higher risk of MASLD and MASLD severity, though the specific diets and more granular assessment of dietary assessment vary across studies. Through this review, we provide further detail regarding the comparative effect of degrees of diet quality and different forms of dietary patterns. Diet quality is challenging to assess but was most commonly defined by the Healthy Eating Index tool which measures diet quality that assesses how well a set of food aligns with key recommendations and dietary patterns published in the Dietary Guidelines for Americans.[47] We also provide comparative analysis of other less commonly studied structural and SDOH beyond race and ethnicity. Results regarding the impact of education across disease prevalence and severity were conflicting, though taken together, lower degrees of formal education appear to have an association with MASH prevalence. The impact of income was highly variable across studies with about half finding significant associations on multivariate analysis, except for clinical outcomes wherein income appeared to be a consistent moderator of risk. This variability is likely a reflection of heterogeneity of variable definition and methods of analysis with income analyzed using multiple parameters including income to poverty ratio, reported income quartiles and other definitions of affluence. Data on the association of insurance type, employment and marital status and MASLD outcomes did not show a significant impact in most studies. In contrast, foreign born status did have a signal with relationship to MASLD disease prevalence and severity but was evaluated in only a small number of studies.

### Implications for clinical care and research

The presence of health disparities in MASLD has been well documented in the literature with significant differences in disease prevalence and severity documented across race, ethnicity and sex.[22] While identification of these disparities is an important first step to identify at-risk populations, identifying the underlying causative factors driving these disparities is necessary to design targeted interventions to move towards health equity. SDOH are hypothesized as being underlying drivers of racial/ethnic disparities in chronic disease including MASLD. Evaluation of SDOH is challenging and complex given their interconnectedness and given that many of these factors are not routinely evaluated in clinical practice. Through this meta-analysis, diet quality appeared to be the most consistent factor associated with disease prevalence and severity, highlighting a priority area for targeted intervention amongst the numerous SDOH factors of interest. From a clinical outcomes perspective, low income demonstrated the strongest association, outlining at-risk populations to which tailored screening and public health interventions would be highest yield.

### Strengths and limitations

There are several limitations to note for this study related to the characteristics and quantity of the existing literature on the topic. A primary limitation stems from the lack of diversified patient populations in which SDOH have been systematically evaluated in MASLD. The majority of studies were conducted in NHANES which limits generalizability to the broader population of patients with MASLD. As outlined in the quality assessment, there are also limitations regarding study design, with the majority of studies being retrospective or cross-sectional analyses which inherently limits the ability to assess for causation. Another limitation of note results from the heterogeneity of definition of outcome across studies. From a meta-analysis standpoint, few SDOH factors were evaluated across studies in a consistent manner, thereby limiting the number of variables that were amenable to pooled meta-analysis. Future studies that adopt the NIMHD recommended assessment strategy for SDOH, PhenX, among prospectively assessed cohorts would greatly strengthen the quality of data and therefore the strength of association found between individual SDOH factors and MASLD outcomes. This in turn can better inform solutions to health equity challenges in MASLD.

## Conclusions

Health disparities in MASLD are an important and unfortunately growing challenge. In this systematic review and meta-analysis, we identified 18 studies including 547,634 individuals in which SDOH were evaluated for their association with prevalence of MASLD, MASH, MASH-associated advanced fibrosis/cirrhosis and clinical outcomes. Beyond race and ethnicity, diet quality, physical inactivity, education and income were the most consistent SDOH factors associated with outcomes of interest. Public health interventions targeted on improving access to healthy eating, particularly among low-income populations appear to be the highest yield area for targeted interventions in MASLD based on the available literature. Future prospective studies employing consistent measures of SDOH are needed to better elucidate the relationship between SDOH and MASLD.

Table 1: Studies Evaluating SDOH Factors and Association with MASH Prevalence

Table 2: Studies Evaluating SDOH Factors and Association with MASH Prevalence

Table 3: Studies Evaluating SDOH Factors and Association with MASH-Associated Advanced Fibrosis/Cirrhosis

## Data Availability

Study materials and data are available in published articles for this meta-analysis.

