## Supplement for "Social Determinants of Health Factors Associated with Metabolic Dysfunction-Associated Steatotic Liver Disease Prevalence and Severity: A Systematic Review and Meta-analysis"

**Supplementary file 1: Specific search strategy**

MEDLINE:

1879 records were identified.

464 records were first excluded (done on animals)

174 records were second excluded (done on pediatric population)

163 records were third excluded (published before 2010)

1079 records matched initial inclusion criteria

((((((("Non-alcoholic Fatty Liver Disease"[Mesh Terms] OR (non-alcoholic fatty liver disease[all fields]) OR (non alcoholic fatty liver disease[all fields])) OR (fatty liver disease[all fields] OR fatty liver disease[Mesh Terms])) OR (Steatohepatitis[all fields] OR Steatohepatitis[Mesh Terms])) OR (Steatonecrosis[all fields] OR Steatonecrosis[Mesh Terms])) OR (Nonalcoholic steatohepatitis[Mesh Terms] OR Nonalcoholic steatohepatitis[all fields])) OR ((metabolic dysfunction-associated steatotic liver disease[Mesh Terms]) OR (metabolic dysfunction-associated steatotic liver disease[all fields]))) OR (MASLD[Mesh Terms] OR MASLD[all fields] OR NAFLD[Mesh Terms] OR NAFLD[all fields])) AND (((((((((((((((((((((((((((((((((((((((Socioeconomic disparities[MeSH Terms])) OR (Underserved populations[MeSH Terms])) OR (Social determinants of health[MeSH Terms])) OR (Income[MeSH Terms])) OR (Education[MeSH Terms])) OR (Cost[MeSH Terms])) OR (food insecurity[MeSH Terms])) OR (Nutrition[MeSH Terms])) OR (housing[MeSH Terms])) OR (Universal coverage[MeSH Terms])) OR (Universal healthcare[MeSH Terms])) OR (Socioeconomic status[MeSH Terms])) OR (Health Inequity[MeSH Terms])) OR (health, urban[MeSH Terms])) OR (vulnerable population[MeSH Terms])) OR (social deprivation index [MeSH Terms])) OR (disparities[Other term])) OR (Underserved populations[Other term])) OR (Social determinants of health[Other term])) OR (Racial disparities[Other term])) OR (Income[Other term])) OR (Education[Other term])) OR (Distance[Other term])) OR (Cost[Other term])) OR (food insecurity[Other term])) OR (urban[Other term])) OR (vulnerable population[Other term])) OR (Nutrition[Other term])) OR (housing[Other term])) OR (Universal health coverage[Other term])) OR (Socioeconomic position[Other term])) OR (Socioeconomic status[Other term])) OR (Inequities[Other term])) OR (Equity[Other term])) OR (inequalities[Other term])) OR (healthcare access[Other term])) OR (Public health[other term])) OR (social deprivation index))

EMBASE:

2547 records were identified.

276 records were first excluded due to duplication. (87 within the same results, 189 with MEDLINE)

179 records were second excluded (done on animals)

45 records were third excluded (Written in language other than English)

165 records were fourth excluded (done on pediatric population)

69 records were fifth excluded (published before 2010)

1813 records matched initial inclusion criteria

((Non-alcoholic Fatty Liver Disease) OR (fatty liver disease) OR Steatohepatitis OR Steatonecrosis OR (Nonalcoholic steatohepatitis) OR (metabolic dysfunction-associated steatotic liver disease) OR MASLD OR NAFLD) AND ((Socioeconomic disparities) OR (Underserved populations) OR (Social determinants of health) OR Income OR (food insecurity) OR housing OR (Universal coverage) OR (Universal healthcare) OR (Socioeconomic status) OR (Health Inequity) OR (urban health) OR (vulnerable population) OR (social deprivation index) OR disparities OR (Racial disparities) OR Distance OR urban OR (Universal health coverage) OR (Socioeconomic position) OR Inequities OR Equity OR inequalities OR (healthcare access))

Cochrane:

338 records were identified.

30 records were protocols

132 records were trials

16 records were third excluded (published before 2010)

160 records matched initial inclusion criteria

((Non-alcoholic Fatty Liver Disease) OR (fatty liver disease) OR (Steatohepatitis) OR (Steatonecrosis) OR (Nonalcoholic steatohepatitis) OR (metabolic dysfunction-associated steatotic liver disease) OR (MASLD) OR (NAFLD)) AND ((Socioeconomic disparities) OR (Underserved populations) OR (Social determinants of health) OR (Income) OR (food insecurity) OR (housing) OR (Universal coverage) OR (Universal healthcare) OR (Socioeconomic status) OR (Health Inequity) OR (urban health) OR (vulnerable population) OR (social deprivation index) OR (disparities) OR (Racial disparities) OR (Distance) OR (urban) OR (Universal health coverage) OR (Socioeconomic position) OR (Inequities) OR (Equity) OR (inequalities) OR (healthcare access))

**Supplement Table 1. Studies Evaluating SDOH Factors and Association with Clinical Outcomes in MASLD/MASH**

| **Author**  **(Year)** | **Study Design** | **Study**  **Cohort**  **N=** | **Clinical Outcomes Assessed** | **SDOH Variables** | **OR or HR (95% CI)** |
| --- | --- | --- | --- | --- | --- |
| Chen  (2023) | Retrospective cohort | UFM cohort, n=15904 | 1- Mortality  2- Cardiovascular disease (CVD)  3- Liver-related events (LRE) | Disadvantage cohort  1- Disadvantage (highest vs lowest) & Mortality  2- Disadvantage (highest vs lowest) & LRE  3- Disadvantage (highest vs lowest) & CVD  4- Black vs white race & Mortality  5- Black vs white race & LRE  6- Black vs white race & CVD  7- Asian vs white race & Mortality  8- Asian vs white race & LRE  9- Asian vs white race & CVD  10- Hispanic vs white race & Mortality  11- Hispanic vs white race & LRE  12- Hispanic vs white race & CVD  13- Other vs white race & Mortality  14- Other vs white race & LRE  15- Other vs white race & CVD  Affluence Cohort  1- Affluence (highest vs lowest) & Mortality  2- Affluence (highest vs lowest) & LRE  3- Affluence (highest vs lowest) & CVD  4- Black vs white race & Mortality  5- Black vs white race & LRE  6- Black vs white race & CVD  7- Asian vs white race & Mortality  8- Asian vs white race & LRE  9- Asian vs white race & CVD  10- Hispanic vs white race & Mortality  11- Hispanic vs white race & LRE  12- Hispanic vs white race & CVD  13- Other vs white race & Mortality  14- Other vs white race & LRE  15- Other vs white race & CVD | 1- aHR 2.08 (1.54–2.81, **p < 0.0001**)  2- aHR 1.36 (0.90–2.07, p = 0.15)  3- aHR 1.36 (1.10–1.68**, p = 0.004)**  4- aHR 0.81 (0.57-1.15, p = 0.24)  5- aHR 0.96 (0.59-1.57, p = 0.88)  6- aHR 0.91 (0.70-1.18, p = 0.46)  7- aHR 0.24 (0.11-0.55**, p = 0.001**)  8- aHR 0.22 (0.07-0.71**, p = 0.01)**  9- aHR 0.52 (0.35-0.77, **p = 0.001)**  10- aHR 0.59 (0.29-1.20, p = 0.14)  11- aHR 0.24 (0.06-0.99, **p = 0.04)**  12- aHR 0.61 (0.37-1.02, p = 0.05)  13- aHR 0.63 (0.31-1.27, p = 0.19)  14- aHR 0.50 (0.16-1.57, p = 0.24)  15- aHR 1.28 (0.87-1.90, p = 0.21)  1- aHR 0.49 (0.37-0.66**, p < 0.0001)**  2- aHR 0.60 (0.39-0.92, **p = 0.02)**  3- aHR 0.71 (0.57–0.88), **p = 0.001)**  4- aHR 0.83 (0.59-1.17, p = 0.29)  5- aHR 1.00 (0.62-1.61, p = 1.0)  6- aHR 0.92 (0.71-1.20, p = 0.55)  7- aHR 0.29 (0.13-0.65**, p = 0.003)**  8- aHR 0.26 (0.08-0.84, **p = 0.02)**  9- aHR 0.56 (0.38-0.84**, p = 0.005)**  10- aHR 0.64 (0.32-1.29, p = 0.21)  11- aHR 0.25 (0.06-1.03, p = 0.05)  12- aHR 0.64 (0.38-1.05, p = 0.07)  13- aHR 0.67 (0.33-1.35, p = 0.26)  14- aHR 0.53 (0.17-1.66, p = 0.27)  15- aHR 1.31 (0.88-1.94, p = 0.18) |
| Kardashian (2022) | Cross-sectional | NHANES (1999-2014), n=34134 | 1- Mortality  2- Healthcare utilization | MASLD cohort and Mortality  1- Food insecurity  2- NH Black vs white  3- Mexican American vs White  4- Other races vs White  5- Income (P:I <1)  6- Insurance (Public vs Private)  7- Insurance (None vs Private)  8- Education (>HS vs <HS)  9- Smoking history (yes vs no)  AF cohort and Mortality  1- Food insecurity  2- NH Black vs white  3- Mexican American vs White  4- Other races vs White  5- Income (P:I <1)  6- Insurance (Public vs Private)  7- Insurance (None vs Private)  8- Education (>HS vs <HS)  9- Smoking history (yes vs no)  MASLD cohort and inpatient health care utilization  1- Food insecurity  2- Income (P:I <1)  3- NH Black vs white  4- Mexican American vs White  5- Other races vs White  6- Insurance (Public vs Private)  7- Insurance (None vs Private)  MASLD cohort and outpatient health care utilization  1- Food insecurity  2- Income (P:I <1)  3- NH Black vs white  4- Mexican American vs White  5- Other races vs White  6- Insurance (Public vs Private)  7- Insurance (None vs Private)  AF cohort and inpatient health care utilization  1- Food insecurity  2- Income (P:I <1)  3- Insurance (Public vs Private)  4- Insurance (None vs Private)  AF cohort and outpatient health care utilization  1- Food insecurity  2- Income (P:I <1)  3- Insurance (Public vs Private)  4- Insurance (None vs Private) | 1- aHR 1.46 (1.08-1.97, **p=0.01)**  2- aHR 1.20 (0.85-1.70, p=0.30)  3- aHR 0.87 (0.66-1.15, p=0.33)  4- aHR 1.07 (0.67-1.73, p=0.77)  5- aHR 1.26 (0.93-1.72, p=0.14)  6- aHR 1.36 (1.06-1.73, **p=0.02)**  7- aHR 0.98 (0.62-1.54, p=0.93)  8- aHR 0.92 (0.70-1.21, p=0.57)  9- aHR 1.45 (1.16-1.81, **p=0.001)**  1- aHR 1.37 (1.01-1.86, **p=0.04)**  2- aHR 0.82 (0.62-1.09, p=0.17)  3- aHR 0.67 (0.45-1.01, p=0.06)  4- aHR 0.75 (0.42-1.34, p=0.33)  5- aHR 0.85 (0.43-1.00**, p=0.05)**  6- aHR 1.27 (1.02-1.57**, p=0.03)**  7- aHR 1.26 (0.71-2.24, p=0.44)  8- aHR 0.70 (0.54-0.91, **p=0.009)**  9- aHR 1.16 (0.93-1.46, p=0.19)  1- aHR 1.46 (0.91-2.34, p=0.12)  2- aHR 1.43 (0.82-2.47, p=0.2)  3- aHR 1.11 (0.62-1.99, p=0.54)  4- aHR 0.76 (0.45-1.27, p=0.29)  5- aHR 0.91 (0.47-1.73, p=0.77)  6- aHR 1.99 (1.38-2.85, **p<0.001)**  7- aHR 1.53 (0.81-2.92, p=0.91)  1- aHR 1.31 (1.05-1.63, **p=0.02)**  2- aHR 1.03 (0.81-1.32, p=0.84)  3- aHR 0.80 (0.63-1.02, p=0.07)  4- aHR 0.53 (0.42-0.65**, p<0.001)**  5- aHR 0.68 (0.51-0.91**, p=0.01)**  6- aHR 1.57 (1.27-1.93, **p<0.001)**  7- aHR 0.42 (0.32-0.54**, p<0.001)**  1- aHR 1.53 (0.83-2.81, p=0.17)  2- aHR 1.24 (0.73-2.11, p=0.43)  3- aHR 1.10 (0.65-1.85, p=0.72)  4- aHR 0.83 (0.31-2.19, p=0.70)  1- aHR 1.09 (0.72-1.66, p=0.68)  2- aHR 1.31 (0.88-1.93, p=0.18)  3- aHR 0.99 (0.75-1.32, p=0.96)  4- aHR 0.31 (0.18-0.53**, p<0.001)** |
| Ruiz-Casas (2021) | Cross-sectional | GAIN Study, n=989 | Fibrosis progression | 1- Smoking (Ex-smoker vs never)  2- Smoking (Current vs never)  3- Employment (part-time vs full-time)  4- Employment (Self-employed vs full-time)  5- Employment (student vs full-time)  6- Employment (retired vs full-time)  7- Employment (unemployed vs full-time)  8- Employment (home-maker vs full-time) | 1- maOR 1.20 (0.74–1.96, p=0.45)  2- maOR 2.01 (1.09–3.66, **p=0.02)**  3- maOR 1.80 (1.02–3.13**, p=0.03)**  4- maOR 0.91 (0.40–1.94, p=0.82)  5- maOR 1.93 (0.39–7.08, p =0.36)  6- maOR 2.22 (1.07–4.51**, p =0.03)**  7- maOR 2.32 (1.12–4.66**, p =0.02)**  8- maOR 1.96 (0.73–4.79, p = 0.15) |
| Yoo  )2020) | Cross-sectional | NHANES (1988-1994), n=10858 | All-cause mortality, cardiovascular disease, and cancer related morality | All-cause mortality  1- Diet quality (high vs poor) (Entire cohort)  2- Diet quality (high vs poor) (Patients with MASLD)  3- Diet quality (high vs poor) (Patients without MASLD)  Cardiovascular disease  1- Diet quality (high vs poor) (Entire cohort)  2- Diet quality (high vs poor) (Patients with MASLD)  3- Diet quality (high vs poor) (Patients without MASLD)  Cancer related mortality  1- Diet quality (high vs poor) (Entire cohort)  2- Diet quality (high vs poor) (Patients with MASLD)  3- Diet quality (high vs poor) (Patients without MASLD) | 1- maOR 0.81 (0.71-0.92**, p=0.002)**  2- maOR 0.98 (0.77-1.10, p=0.48)  3- maOR 0.91 (0.86-0.96, **p=0.002)**  1- maOR 0.90 (0.64-1.28, p=0.55)  2- maOR 0.97 (0.54-1.72, p=0.90)  2- maOR 0.86 (0.60-1.24, p=0.41)  1- maOR 0.64 (0.48-0.85, **p=0.003)**  2- maOR 0.52 (0.35-0.80**, p=0.004)**  3- maOR 0.88 (0.54-1.42, p=0.58) |
| Le  (2020) | Cross-sectional | NHANES (2011-2016), n=4538 | Health care utilization, awareness of liver disease | Received healthcare 1-3 times in the past year  1- Foreign born  2- Insured  3- Income to poverty ratio  Received healthcare 4-9 times in the past year  1- Foreign born  2- Insured  3- Income to poverty ratio  Received healthcare +10 times in the past year  1- Foreign born  2- Insured  3- Income to poverty ratio  Aware of liver disease  Healthcare service utilization | 1- maOR 1.03 (0.60-1.78, p=0.91)  2- maOR 3.70 (1.81-7.58**, p=0.001)**  3- maOR 1.38 (1.10-1.74, **p=0.007)**  1- maOR 0.56 (0.34-0.93**, p=0.02)**  2- maOR 5.38 (2.07-13.93**, p=0.001)**  3- maOR 1.05 (0.79-1.39, p=0.72)  1- maOR 0.41 (0.21-0.83, **p=0.01)**  2- maOR 12.64 (4.00-39.9**, p<0.001)**  3- maOR 1.00 (0.71-1.41, p=0.99)  maOR 1.54 (1.01-2.34**, p=0.04)** |
| Adejumo (2019) | Retrospective cohort | National Inpatient Sample (2007-2014), n=210660 | In hospital mortality, discharge disposition, hospital charges, length of stay | In hospital mortality  1- Race (black vs white)  2- Race (Hispanic vs white)  3- Race (Asians & others vs white)  4- Health insurance (Government vs Private)  5- Health insurance (Self pay and others vs Private)  6- Income status (Lowest vs highest quartile)  7- Hospital region (Midwest vs northeast)  8- Hospital region (South vs northeast)  9- Hospital region (West vs northeast)  10- Hospital teaching status (Urban nonteaching vs rural)  11- Hospital teaching status (Urban teaching vs rural)  12- Liver cirrhosis (compensated vs no-cirrhosis)  13- Liver cirrhosis (decompensated vs no-cirrhosis)  Discharge disposition  1- Race (black vs white)  2- Race (Hispanic vs white)  3- Race (Asians & others vs white)  4- Health insurance (Government vs Private)  5- Health insurance (Self pay and others vs Private)  6- Income status (Lowest vs highest quartile)  7- Hospital region (Midwest vs northeast)  8- Hospital region (South vs northeast)  9- Hospital region (West vs northeast)  10- Hospital teaching status (Urban nonteaching vs rural)  11- Hospital teaching status (Urban teaching vs rural)  12- Liver cirrhosis (compensated vs no-cirrhosis)  13- Liver cirrhosis (decompensated vs no-cirrhosis)  Hospital charges  1- Race (black vs white)  2- Race (Hispanic vs white)  3- Race (Asians & others vs white)  4- Health insurance (Government vs Private)  5- Health insurance (Self pay and others vs Private)  6- Income status (Lowest vs highest quartile)  7- Hospital region (Midwest vs northeast)  8- Hospital region (South vs northeast)  9- Hospital region (West vs northeast)  10- Hospital teaching status (Urban nonteaching vs rural)  11- Hospital teaching status (Urban teaching vs rural)  12- Liver cirrhosis (compensated vs no-cirrhosis)  13- Liver cirrhosis (decompensated vs no-cirrhosis)  Length of stay  1- Race (black vs white)  2- Race (Hispanic vs white)  3- Race (Asians & others vs white)  4- Health insurance (Government vs Private)  5- Health insurance (Self pay and others vs Private)  6- Income status (Lowest vs highest quartile)  7- Hospital region (Midwest vs northeast)  8- Hospital region (South vs northeast)  9- Hospital region (West vs northeast)  10- Hospital teaching status (Urban nonteaching vs rural)  11- Hospital teaching status (Urban teaching vs rural)  12- Liver cirrhosis (compensated vs no-cirrhosis)  13- Liver cirrhosis (decompensated vs no-cirrhosis) | In hospital mortality  1- maOR 1.02 (0.83-1.25, p=1.00)  2- maOR 0.84 (0.69-1.03, p=0.13)  3- maOR 0.99 (0.78-1.25, p=1.00)  4- maOR 1.02 (1.00-1.30, p=0.06)  5- maOR 1.14 (1.09-1**.67, p=0.002)**  6- maOR 1.18 (0.99-1.14, p=0.08)  7- maOR 1.18 (0.96-1.45, p=0.22)  8- maOR 1.22 (1.01-1.48, **p=0.02)**  9- maOR 1.41 (1.15-1.73, **p<0.0001)**  10- maOR 1.16 (0.93-1.44, p=0.33)  11- maOR 1.42 (1.14-1.76**, p=0.0003)**  12- maOR 2.34 (1.93-2.84, **p<0.0001)**  13- maOR 7.56 (6.15-9.28, **p<0.0001)**  Unfavorable discharge disposition  1- maOR 1.14 (1.06-1.22, **p<0.0001)**  2- maOR 0.71 (0.66-0.76, **p<0.0001)**  3- maOR 0.89 (0.81-0.98**, p=0.0082)**  4- maOR 1.90 (1.81-2.00, **p<0.0001)**  5- maOR 1.24 (1.14-1.34**, p<0.0001)**  6- maOR 1.04 (0.97-1.12, p=0.82)  7- maOR 0.99 (0.91-1.09, p>0.99)  8- maOR 0.80 (0.74-0.87, **p<0.0001)**  9- maOR 0.90 (0.83-0.98**, p=0.010)**  10- maOR 0.90 (0.93-0.97, **p=0.003)**  11- maOR 0.83 (0.77-0.90**, p<0.0001)**  12- maOR 1.18 (1.07-1.30, **p=0.0001)**  13- maOR 1.19 (1.09-1.31**, p<0.0001)**  Hospital charges  1- maOR 1.00 (0.94-1.07, p=1.00)  2- maOR 1.09 (1.04-1.14, **p<0.0001)**  3- maOR 1.09 (1.03-1.15, **p=0.0005)**  4- maOR 1.01 (0.99-1.03, p=1.00)  5- maOR 0.97 (0.94-1.00**, p=0.03)**  6- maOR 0.98 (0.93-1.04, p=1.00)  7- maOR 0.90 (0.81-0.99**, p=0.02)**  8- maOR 1.00 (0.91-1.10, p=1.00)  9- maOR 1.42 (1.29-1.56, **p<0.0001)**  10- maOR 1.59 (1.49-1.70, **p<0.0001)**  11- maOR 1.83 (1.70-1.96, **p<0.0001)**  12- maOR 0.97 (0.90-1.03, p=0.58)  13- maOR 0.76 (0.72-0.81, **p<0.0001)**  Length of stay  1- maOR 1.07 (1.04-1.10**, p<0.0001)**  2- maOR 0.98 (0.95-1.00, p=0.11)  3- maOR 1.02 (0.99-1.06, p=0.43)  4- maOR 1.19 (1.17-1.22**, p<0.0001)**  5- maOR 1.10 (1.06-1.13**, p<0.0001)**  6- maOR 1.04 (1.01-1.07, **p=0.002)**  7- maOR 0.95 (0.91-1.00, p=0.03)  8- maOR 0.99 (0.95-1.03, p>0.99)  9- maOR 0.93 (0.89-0.98, **p=0.0009)**  10- maOR 1.13 (1.09-1.17**, p<0.0001)**  11- maOR 1.23 (1.19-1.27, **p<0.0001)**  12- maOR 0.97 (0.92-1.01, p=0.17)  13- maOR 0.83 (0.79-0.86, **p<0.0001)** |
| Alvi  (2023) | Retrospective cohort | National Inpatient Sample (2019), n=196860 | CVD and mortality | High vs low-income quartile cohorts  1- All-cause mortality  2- Acute myocardial infarction  3- Cardiac arrest  4- Acute PE  5- Stroke | 1- maOR 0.77 (0.62-0.96**, p=0.022)**  2- maOR 0.78 (0.65-0.93**, p=0.005)**  3- maOR p=0.23  4- maOR p=0.28  5- maOR p=0.29 |
| Chen  (2023) | Retrospective cohort | NHANES 1988-1994 (n=9068) and NHANES 1999-2014 (n=14963) | Mortality | Physical activity (Active vs inactive) | 1- maOR 0.77 (0.60–0.99, **p=0.04)** |
| Han  (2021) | Cross-sectional | Tertiary academic center | Disease severity | This is baseline characteristics (they compared patients with satisfactory vs unsatisfactory responses and measured their LSM score), they did not do any multivariate analysis to see if any covariate predicted | No differences were found between both cohorts (p values >0.05). |

SDOH: social determinant of health; MASLD: metabolic dysfunction-associated steatotic liver disease; OR: odds ratio; NHANES: National Health and Nutrition Examination Survey; HS: high school; aaOR: Age-adjusted odds ratio; maOR: Multivariate-adjusted odds ratio; NH: Non-Hispanic; P:I : Poverty to income ratio; NH: non-Hispanic; LRE: liver related event; PE: pulmonary embolism; CVD: cardiovascular disease;

**Supplement Table 2: Association between Individual SDOH Factors on MASLD/MASH**

| **SDOH** | **Outcome** | **Studies predicting increased risk** | **Studies predicting decreased risk** | **Studies predicting similar risk** |
| --- | --- | --- | --- | --- |
| **Black race (vs white)** | MASLD prevalence |  | Unalp-Arida A (2023)  maOR 0.51 (0.39–0.65, p<0.001) | Black race only  Giammarino (2022)  uaOR 1.00 (0.94‐1.00, p=0.97) |
|  |  |  | Golovaty (2020)  maOR 0.42 (0.31-0.58, p<0.001) |  |
|  |  |  | Le (2020)  maOR 0.44 (0.31-0.62, p<0.001) |  |
|  | MASH prevalence |  |  | Black race only  Giammarino (2022)  uaOR 0.99 (0.98‐1.00, p=0.20) |
|  | Advanced Cirrhosis and Fibrosis prevalence |  |  | Unalp-Arida A  (2023)  maOR 0.78 (0.53-1.15, p=0.20) |
|  |  |  |  | Golovaty (2020)  maOR 1.00 (0.60-1.67, p>0.05) |
|  |  |  |  | Le (2020)  maOR 1.43 (0.85-2.39, p=0.17) |
|  |  |  |  | Black race only  Giammarino (2022)  uaOR 0.99 (0.99‐1.00, p=0.73) |
|  | All-cause mortality |  |  | Chen VL (2023) (Disadvantage cohort)  aHR 0.81 (0.57-1.15, p = 0.24) |
|  |  |  |  | Chen VL (2023) (Affluence cohort)  aHR 0.83 (0.59-1.17, p = 0.29) |
|  |  |  |  | Kardashian A (2022) (NAFLD Cohort)  aHR 1.20 (0.85-1.70, p=0.30) |
|  |  |  |  | Kardashian A (2022) (AF Cohort)  aHR 0.82 (0.62-1.09, p=0.17) |
|  |  |  |  | Adejumo AC (2019) (IH)  maOR 1.02 (0.83-1.25, p=1.00) |
|  | Cardiovascular disease |  |  | Chen VL (2023) (Disadvantage cohort)  asHR 0.91 (0.70-1.18, p = 0.46) |
|  |  |  |  | Chen VL (2023) (Affluence cohort)  asHR 0.92 (0.71-1.20, p = 0.55) |
|  | Liver related events |  |  | Chen VL (2023) (Disadvantage cohort)  asHR 0.96 (0.59-1.57, p = 0.88) |
|  |  |  |  | Chen VL (2023) (Affluence cohort)  asHR 1.00 (0.62-1.61, p = 1.0) |
|  | Inpatient healthcare utilization |  |  | Kardashian A (2022) (MASLD Cohort)  aHR 1.11 (0.62-1.99, p=0.54) |
|  | Outpatient healthcare utilization |  |  | Kardashian A (2022) (MASLD Cohort)  aHR 0.80 (0.63-1.02, p=0.07) |
|  | Unfavorable discharge disposition | Adejumo AC (2019)  maOR 1.14 (1.06-1.22, p<0.0001) |  |  |
|  | Hospital charges |  |  | Adejumo AC (2019)  maOR 1.00 (0.94-1.07, p=1.00) |
|  | Increased length of stay | Adejumo AC (2019)  maOR 1.07 (1.04-1.10, p<0.0001) |  |  |

| **SDOH** | **Outcome** | **Studies predicting increased risk** | **Studies predicting decreased risk** | **Studies predicting similar risk** |
| --- | --- | --- | --- | --- |
| **Asian race (vs white)** | MASLD prevalence | Unalp-Arida A (2023)  maOR 1.49 (1.05–2.10, p=0.02) |  | Le (2020)  maOR 0.95 (0.51-1.77, p=0.87) |
|  | Advanced Cirrhosis and Fibrosis prevalence |  | Le (2020)  maOR 0.12 (0.29-0.50, p=0.005) | Unalp-Arida A  (2023)  maOR 1.21 (0.77-1.90, p=0.40) |
|  | All-cause mortality |  | Chen VL (2023) (Disadvantage cohort)  aHR 0.24 (0.11-0.55, p = 0.001) | Adejumo AC (2019) (IH)  maOR 0.99 (0.78-1.25, p=1.00) |
|  |  |  | Chen VL (2023) (Affluence cohort)  aHR 0.29 (0.13-0.65, p = 0.003) |  |
|  | Cardiovascular disease |  | Chen VL (2023) (Disadvantage cohort)  asHR 0.52 (0.35-0.77, p = 0.001) |  |
|  |  |  | Chen VL (2023) (Affluence cohort)  asHR 0.56 (0.38-0.84, p = 0.005) |  |
|  | Liver related events |  | Chen VL (2023) (Disadvantage cohort)  asHR 0.22 (0.07-0.71, p = 0.01) |  |
|  |  |  | Chen VL (2023) (Affluence cohort)  asHR 0.26 (0.08-0.84, p = 0.02) |  |
|  | Unfavorable discharge disposition |  | Adejumo AC (2019)  maOR 0.89 (0.81-0.98, p=0.008) |  |
|  | Hospital charges | Adejumo AC (2019)  maOR 1.09 (1.03-1.15, p=0.0005) |  |  |
|  | Increased length of stay |  |  | Adejumo AC (2019)  maOR 1.02 (0.99-1.06, p=0.43) |

| **SDOH** | **Outcome** | **Studies predicting increased risk** | **Studies predicting decreased risk** | **Studies predicting similar risk** |
| --- | --- | --- | --- | --- |
| **Non-Mexican Hispanic (vs white)** | MASLD prevalence |  |  | Unalp-Arida A (2023)  maOR 1.17 (0.86–1.59, p=0.31) |
|  |  |  |  | Golovaty (2020)  maOR 0.93 (0.68-1.27, p>0.05) |
|  |  |  |  | Le (2020)  maOR 1.41 (0.79-2.50, p=0.23) |
|  |  |  |  | Hispanic race only  Giammarino (2022)  uaOR 1.00 (0.99‐1.01, p=0.20) |
|  | MASH prevalence |  |  | Bambha K (2012)  maOR 1.43 (0.88-2.33, p=0.15) |
|  |  |  |  | Hispanic race only  Giammarino (2022)  uaOR 0.99 (0.98‐1.01, p=0.72) |
|  | Advanced Cirrhosis and Fibrosis prevalence |  | Golovaty (2020)  maOR 0.32 (0.16, 0.66, p<0.05) | Unalp-Arida A (2023)  maOR 0.94 (0.65-1.35, p=0.72) |
|  |  |  |  | Bambha K (2012)  maOR 0.78 (0.42-1.45, p=0.43) |
|  |  |  |  | Le (2020)  maOR 1.03 (0.46-2.28, p=0.94) |
|  |  |  |  | Hispanic race only  Giammarino (2022)  uaOR 1.00 (0.99‐1.01, p=0.13) |
|  |  |  |  | Rieman-Klinger (2024)  maOR 1.79 (0.63–5.07, p=0.27) |
|  | All-cause mortality |  |  | Chen VL (2023) (Disadvantage cohort)  aHR 0.59 (0.29-1.20, p = 0.14) |
|  |  |  |  | Chen VL (2023) (Affluence cohort)  aHR 0.64 (0.32-1.29, p = 0.21) |
|  |  |  |  | Adejumo AC (2019) (IH)  maOR 0.84 (0.69-1.03, p=0.13) |
|  | Cardiovascular disease |  |  | Chen VL (2023) (Disadvantage cohort)  asHR 0.61 (0.37-1.02, p = 0.06) |
|  |  |  |  | Chen VL (2023) (Affluence cohort)  asHR 0.64 (0.38-1.05, p = 0.07) |
|  | Liver related events |  | Chen VL (2023) (Disadvantage cohort)  asHR 0.24 (0.06-0.99, p = 0.04) | Chen VL (2023) (Affluence cohort)  asHR 0.25 (0.06-1.03, p = 0.06) |
|  | Unfavorable discharge disposition |  | Adejumo AC (2019)  maOR 0.71 (0.66-0.76, p<0.0001) |  |
|  | Hospital charges | Adejumo AC (2019)  maOR 1.09 (1.04-1.14, p<0.0001) |  |  |
|  | Increased length of stay |  |  | Adejumo AC (2019)  maOR 0.98 (0.95-1.00, p=0.11) |

| **SDOH** | **Outcome** | **Studies predicting increased risk** | **Studies predicting decreased risk** | **Studies predicting similar risk** |
| --- | --- | --- | --- | --- |
| **Mexican race (vs white)** | MASLD prevalence | Le (2020)  maOR 3.09 (2.07-4.60, p<0.001) |  |  |
|  |  | Golovaty (2020)  maOR 2.03 (1.50, 2.74, p<0.001) |  |  |
|  | Advanced Cirrhosis and Fibrosis prevalence |  | Le (2020)  maOR 0.40 (0.24-0.67, p=0.001) | Golovaty (2020)  maOR 1.46 (0.78, 2.70, p>0.05) |
|  | All-cause mortality |  |  | Kardashian A (2022) (MASLD Cohort)  aHR 0.87 (0.66-1.15, p=0.33) |
|  |  |  |  | Kardashian A (2022) (AF Cohort)  aHR 0.67 (0.45-1.01, p=0.06) |
|  | Inpatient healthcare utilization |  |  | Kardashian A (2022) (MASLD Cohort)  aHR 0.76 (0.45-1.27, p=0.29) |
|  | Outpatient healthcare utilization |  | Kardashian A (2022) (NAFLD Cohort)  aHR 0.53 (0.42-0.65, p<0.001) |  |

| **SDOH** | **Outcome** | **Studies predicting increased risk** | **Studies predicting decreased risk** | **Studies predicting similar risk** |
| --- | --- | --- | --- | --- |
| **Other races (vs white)**  **Includes but not limited to: Asian, Pacific Islander, Native American, Alaskan**  ***Limitation** | MASLD prevalence |  | Golovaty (2020)  maOR 0.50 (0.32, 0.77, p<0.01) | Le (2020)  maOR 1.38 (0.65-2.94, p=0.39) |
|  | Advanced Cirrhosis and Fibrosis prevalence |  |  | Golovaty (2020)  maOR 0.44 (0.16, 1.18, p>0.05) |
|  |  |  |  | Le (2020)  maOR 0.64 (0.35-1.19, p=0.15) |
|  | All-cause mortality |  |  | Chen VL (2023) (Disadvantage cohort)  aHR 0.63 (0.31-1.27, p = 0.19) |
|  |  |  |  | Chen VL (2023) (Affluence cohort)  aHR 0.67 (0.33-1.35, p = 0.26) |
|  |  |  |  | Kardashian A (2022) (NAFLD Cohort)  aHR 1.07 (0.67-1.73, p=0.77) |
|  |  |  |  | Kardashian A (2022) (AF Cohort)  aHR 0.75 (0.42-1.34, p=0.33) |
|  | Cardiovascular disease |  |  | Chen VL (2023) (Disadvantage cohort)  asHR 1.28 (0.87-1.90, p = 0.21) |
|  |  |  |  | Chen VL (2023) (Affluence cohort)  asHR 1.31 (0.88-1.94, p = 0.18) |
|  | Liver related events |  |  | Chen VL (2023) (Disadvantage cohort)  asHR 0.50 (0.16-1.57, p = 0.24) |
|  |  |  |  | Chen VL (2023) (Affluence cohort)  asHR 0.53 (0.17-1.66, p = 0.27) |
|  | Inpatient healthcare utilization |  |  | Kardashian A (2022) (MASLD Cohort)  aHR 0.91 (0.47-1.73, p=0.77) |
|  | Outpatient healthcare utilization |  | Kardashian A (2022) (MASLD Cohort)  aHR 0.68 (0.51-0.91, p=0.01) |  |

| **SDOH** | **Outcome** | **Studies predicting increased risk** | **Studies predicting decreased risk** | **Studies predicting similar risk** |
| --- | --- | --- | --- | --- |
| **Income** | MASLD prevalence | (P:I <1.65 vs >5)  Unalp-Arida A (2023)  aaOR 1.36 (1.02-1.82, p=0.03) |  | (P:I 1.65-3.2 vs >5)  Unalp-Arida A (2023)  aaOR 1.32 (0.94-1.85, p=0.11) |
|  |  |  |  | (<25k vs >25k)  Golovaty (2020)  maOR 1.08 (0.76, 1.55, p>0.05) |
|  |  |  |  | (I:P <1)  Le (2020)  maOR 0.91 (0.82-1.00, p=0.059) |
|  |  |  |  | Poverty  Giammarino (2022)  uaOR 1.005 (0.998‐1.012, p=0.17) |
|  | MASH prevalence |  | (>50k vs <50k)  Bambha K (2012)  uaOR 0.64 (0.49-0.85, p<0.01) | Poverty  Giammarino (2022)  uaOR 1.003 (0.993‐1.013, p=0.52) |
|  | Advanced Cirrhosis and Fibrosis prevalence | (P:I <1.65 vs >5)  Unalp-Arida A (2023)  maOR 2.00 (1.18-3.40, p=0.01) |  | (<25k vs >25k)  Golovaty (2020)  maOR 2.29 (0.99, 5.32, p>0.05) |
|  |  | (P:I 1.65-3.2 vs >5)  Unalp-Arida A (2023)  maOR 2.59 (1.38-4.85, p=0.005) |  | Poverty  Giammarino (2022)  uaOR 1.006 (0.998‐1.015, p=0.14) |
|  |  | (<30k vs >30k)  Rieman-Klinger (2024)  maOR 0.26 (0.08–0.85, p=0.027) |  |  |
|  | All-cause mortality |  | (P:I <1)  Kardashian A (2022) (AF Cohort)  aHR 0.85 (0.43-1.00, p=0.05) | (P:I <1)  Kardashian A (2022) (MASLD Cohort)  aHR 1.26 (0.93-1.72, p=0.14) |
|  |  |  | (Highest vs lowest quartiles)  Alvi (2023)  maOR 0.77 (0.62-0.96, p=0.02) | (Lowest vs highest)  Adejumo AC (2019) (IH)  maOR 1.18 (0.99-1.14, p=0.08) |
|  | Cardiovascular disease mortality |  | (Highest vs lowest quartiles)  Alvi (2023)  maOR 0.78 (0.65-0.93, p=0.005) |  |
|  | Inpatient healthcare utilization |  |  | (P:I <1)  Kardashian A (2022) (MASLD Cohort)  aHR 1.43 (0.82-2.47, p=0.2) |
|  |  |  |  | (P:I <1)  Kardashian A (2022) (AF Cohort)  aHR 1.24 (0.73-2.11, p=0.43) |
|  | Outpatient healthcare utilization |  |  | (P:I <1)  Kardashian A (2022) (MASLD Cohort)  aHR 1.03 (0.81-1.32, p=0.84) |
|  |  |  |  | (P:I <1)  Kardashian A (2022) (AF Cohort)  aHR 1.31 (0.88-1.93, p=0.18) |
|  | Received healthcare 1-3 times in past year | (I:P <1)  Le (2020)  maOR 1.38 (1.10-1.74, p=0.007) |  |  |
|  | Received healthcare 4-9 times in past year |  |  | (I:P <1)  Le (2020)  maOR 1.05 (0.79-1.39, p=0.72) |
|  | Received healthcare +10 times in past year |  |  | (I:P <1)  Le (2020)  maOR 1.00 (0.71-1.41, p=0.99) |
|  | Unfavorable discharge disposition |  |  | Lowest vs highest quartiles  Adejumo AC (2019)  maOR 1.04 (0.97-1.12, p=0.82) |
|  | Hospital charges |  |  | Lowest vs highest quartiles  Adejumo AC (2019)  maOR 0.98 (0.93-1.04, p=1.00) |
|  | Increased length of stay | Lowest vs highest quartiles  Adejumo AC (2019)  maOR 1.04 (1.01-1.07, p=0.002) |  |  |

| **SDOH** | **Outcome** | **Studies predicting increased risk** | **Studies predicting decreased risk** | **Studies predicting similar risk** |
| --- | --- | --- | --- | --- |
| **Education** | MASLD prevalence | (<HS vs >HS)  Unalp-Arida A (2023)  aaOR 1.36 (1.06-1.76, p=0.02) |  | (<HS vs >HS)  Golovaty (2020)  maOR 0.82 (0.62, 1.08, p>0.05) |
|  |  | (HS vs >HS)  Unalp-Arida A (2023)  aaOR 1.58 (1.24,2.01, p<0.001 |  | (HS vs >HS)  Golovaty (2020)  maOR 0.89 (0.67, 1.18, p>0.05) |
|  |  |  |  | (<HS vs >HS)  Le (2020)  maOR 0.80 (0.48-1.36, p=0.40) |
|  |  |  |  | (<HS vs HS)  Le (2020)  maOR 0.96 (0.61-1.52, p=0.86) |
|  |  |  |  | Only high school diploma  Giammarino (2022)  uaOR 0.996 (0.977‐1.016, p=0.69) |
|  |  |  |  | Bachelor’s degree or higher  Giammarino (2022)  uaOR 1.000 (0.990‐1.011, p=0.93) |
|  |  |  |  | High school drop out  Giammarino (2022)  uaOR 1.004 (0.998‐1.009, p=0.19) |
|  | MASH prevalence | Bachelor’s degree or higher  Giammarino (2022)  uaOR 1.016 (1.00‐1.03, p=0.02) | (>HS vs <HS)  Bambha K (2012)  uaOR 0.74 (0.55-1.00, p=0.05) | High school drop out  Giammarino (2022)  uaOR 0.996 (0.988‐1.005, p=0.37)) |
|  |  |  | Only high school diploma  Giammarino (2022)  uaOR 0.969 (0.941‐0.997, p=0.03) |  |
|  | Advanced Cirrhosis and Fibrosis prevalence | (HS vs >HS)  Unalp-Arida A (2023)  maOR 1.77 (1.05-3.00, p=0.034) |  | (<HS vs >HS)  Unalp-Arida A (2023)  maOR 1.29 (0.85-1.96, p=0.22) |
|  |  | (<HS vs >HS)  Rieman-Klinger (2024)  maOR 8.04 (1.9–33.85, p=0.005) |  | (<HS vs >HS)  Golovaty (2020)  maOR 0.70 (0.40, 1.23, p>0.05) |
|  |  |  |  | (HS vs >HS)  Golovaty (2020)  maOR 0.79 (0.43, 1.44, p>0.05) |
|  |  |  |  | Only high school diploma  Giammarino (2022)  uaOR 0.995 (0.97‐1.02, p=0.69) |
|  |  |  |  | Bachelor’s degree or higher  Giammarino (2022)  uaOR 1.001 (0.9‐1.0, p=0.83) |
|  |  |  |  | High school drop out  Giammarino (2022)  uaOR 1.007 (0.9‐1.01, p=0.06) |
|  | All-cause mortality |  | (>HS vs <HS)  Kardashian A (2022) (AF Cohort)  aHR 0.70 (0.54-0.91, p=0.009) | (>HS vs <HS)  Kardashian A (2022) (NAFLD Cohort)  aHR 0.92 (0.70-1.21, p=0.57) |

| **SDOH** | **Outcome** | **Studies predicting increased risk** | **Studies predicting decreased risk** | **Studies predicting similar risk** |
| --- | --- | --- | --- | --- |
| **Insurance**  **(vs private)** | MASLD prevalence |  |  | Public  Unalp-Arida A (2023)  aaOR 1.08 (0.89-1.32, p=0.40) |
|  |  |  |  | Uninsured  Unalp-Arida A (2023)  aaOR 0.97 (0.78-1.21, p=0.78) |
|  |  |  |  | Private healthcare (alone)  Giammarino (2022)  uaOR 0.99 (0.97‐1.00, p=0.14) |
|  |  |  |  | Public healthcare (alone)  Giammarino (2022)  uaOR 1.00 (0.987‐1.028, p=0.50) |
|  |  |  |  | Uninsured (alone)  Giammarino (2022)  uaOR 1.03 (0.99‐1.07, p=0.14) |
|  | MASH prevalence |  |  | Private healthcare (alone)  Giammarino (2022)  uaOR 0.99 (0.98‐1.01, p=0.85) |
|  |  |  |  | Public healthcare (alone)  Giammarino (2022)  uaOR 1.00 (0.975‐1.035, p=0.77) |
|  |  |  |  | Uninsured (alone)  Giammarino (2022)  uaOR 0.99 (0.93‐1.05, p=0.86) |
|  | Advanced Cirrhosis and Fibrosis prevalence | Public healthcare (alone)  Giammarino (2022)  uaOR 1.02 (1.00‐1.05, p=0.03) | Private healthcare (alone)  Giammarino (2022)  uaOR 0.98 (0.96‐0.99, p=0.04) | Public  Unalp-Arida A (2023)  aaOR 1.06 (0.81-1.39, p=0.66) |
|  |  |  |  | Uninsured  Unalp-Arida A (2023)  aaOR 1.04 (0.72-1.50, p=0.84) |
|  |  |  |  | Uninsured (alone)  Giammarino (2022)  uaOR 1.03 90.98‐1.80, p=0.17) |
|  | All-cause mortality | Public  Kardashian A (2022) (MASLD Cohort)  aHR 1.36 (1.06-1.73, p=0.02) |  | Public  Adejumo AC (2019) (IH)  maOR 1.02 (1.00-1.30, p=0.06) |
|  |  | Public  Kardashian A (2022) (AF Cohort)  aHR 1.27 (1.02-1.57, p=0.03) |  | Uninsured  Kardashian A (2022) (MASLD Cohort)  aHR 0.98 (0.62-1.54, p=0.93) |
|  |  | Uninsured  Adejumo AC (2019) (IH)  maOR 1.14 (1.09-1.67, p=0.002) |  | Uninsured  Kardashian A (2022) (MASLD Cohort)  aHR 1.26 (0.71-2.24, p=0.44) |
|  | Inpatient healthcare utilization | Public  Kardashian A (2022) (MASLD Cohort)  aHR 1.99 (1.38-2.85, p<0.001) |  | Public  Kardashian A (2022) (AF Cohort)  aHR 1.10 (0.65-1.85, p=0.72) |
|  |  |  |  | Uninsured  Kardashian A (2022) (MASLD Cohort)  aHR 1.53 (0.81-2.92, p=0.91) |
|  |  |  |  | Uninsured  Kardashian A (2022) (AF Cohort)  aHR 0.83 (0.31-2.19, p=0.70) |
|  | Outpatient healthcare utilization | Public  Kardashian A (2022) (NAFLD Cohort)  aHR 1.57 (1.27-1.93, p<0.001) | Uninsured  Kardashian A (2022) (NAFLD Cohort)  aHR 0.42 (0.32-0.54, p<0.001) | Public  Kardashian A (2022) (AF Cohort)  aHR 0.99 (0.75-1.32, p=0.96) |
|  |  |  | Uninsured  Kardashian A (2022) (AF Cohort)  aHR 0.31 (0.18-0.53, p<0.001) |  |
|  | Received healthcare 1-3 times in past year | Yes vs no  Le (2020)  maOR 3.70 (1.81-7.58, p=0.001) |  |  |
|  | Received healthcare 4-9 times in past year | Yes vs no  Le (2020)  maOR 5.38 (2.07-13.93, p=0.001) |  |  |
|  | Received healthcare +10 times in past year | Yes vs no  Le (2020)  maOR 12.64 (4.00-39.9, p<0.001) |  |  |
|  | Unfavorable discharge disposition | Public  Adejumo AC (2019)  maOR 1.90 (1.81-2.00, p<0.0001) |  |  |
|  |  | Uninsured  Adejumo AC (2019)  maOR 1.24 (1.14-1.34, p<0.0001) |  |  |
|  | Hospital charges |  | Uninsured  Adejumo AC (2019)  maOR 0.97 (0.94-1.00, p=0.03) | Public  Adejumo AC (2019)  maOR 1.01 (0.99-1.03, p=1.00) |
|  | Increased length of stay | Public  Adejumo AC (2019)  maOR 1.19 (1.17-1.22, p<0.0001) |  |  |
|  |  | Uninsured  Adejumo AC (2019)  maOR 1.10 (1.06-1.13, p<0.0001) |  |  |

| **SDOH** | **Outcome** | **Studies predicting increased risk** | **Studies predicting decreased risk** | **Studies predicting similar risk** |
| --- | --- | --- | --- | --- |
| **Smoking** | MASLD prevalence | Former smoker  Unalp-Arida A (2023)  aaOR 1.40 (1.19–1.66, p<0.001) |  | Current smoker  Unalp-Arida A (2023)  aaOR 1.05 (0.83–1.33, p=0.65) |
|  |  |  |  | Either (Current or former)  Golovaty (2020)  maOR 1.04 (0.84, 1.28, p>0.05) |
|  |  |  |  | Either (Current or former)  Le (2020)  maOR 1.09 (0.78-1.51, p=0.60) |
|  | Advanced Cirrhosis and Fibrosis prevalence | Former smoker  Unalp-Arida A (2023)  aaOR 1.30 (1.03-1.64, p=0.02) |  | Current smoker  Unalp-Arida A (2023)  aaOR 1.12 (0.90-1.39, p=0.30) |
|  |  |  |  | Either (Current or former)  Golovaty (2020)  maOR 0.74 (0.46, 1.17, p>0.05) |
|  |  |  |  | Either (Current or former)  Le (2020)  maOR 1.30 (0.85-1.99, p=0.22) |
|  | All-cause mortality | Kardashian A (2022) (NAFLD Cohort)  aHR 1.45 (1.16-1.81, p=0.001) |  | Kardashian A (2022) (AF Cohort)  aHR 1.16 (0.93-1.46, p=0.19) |
|  | Fibrosis progression | Current  Ruiz-Casas L (2021)  maOR 2.01 (1.09–3.66, p=0.02) |  | Former  Ruiz-Casas L (2021)  maOR 1.20 (0.74–1.96, p=0.45) |

| **SDOH** | **Outcome** | **Studies predicting increased risk** | **Studies predicting decreased risk** | **Studies predicting similar risk** |
| --- | --- | --- | --- | --- |
| **Diet quality (high vs poor)** | MASLD prevalence | Fast food (> 7 times / week vs none)  Hu (2022)  maOR 1.70 (1.15-2.51, p=0.01) | (high vs poor)  Yoo ER (2020)  maOR 0.86 (0.77-0.96, p=0.006) | (poor vs excellent)  Unalp-Arida A (2023)  maOR 1.28 (0.97–1.69, p=0.07) |
|  |  |  | DASH (Q5 vs Q1)  Park SY (2020)  maOR 0.80 (0.69-0.92, p=0.005) | AHIE (Q5 vs Q1)  Park SY (2020)  maOR 0.96 (0.84-1.10, p=0.82) |
|  |  |  | HIE (Q5 vs Q1)  Park SY (2020)  maOR 0.84 (0.74-0.97, p=0.03) | AMD (Q5 vs Q1)  Park SY (2020)  maOR 1.00 (0.86-1.15, p=0.63) |
|  |  |  | DASH (High vs low) – FHS  Gao (2023)  maOR 0.68 (0.61-0.76, p<0.05) | DASH (High vs low) – NHANES  Gao (2023)  maOR 0.84 (0.71-1.00, p>0.05) |
|  |  |  | AHEI (High vs low) – FHS  Gao (2023)  maOR 0.66 (0.59-0.73, p<0.05) |  |
|  |  |  | MDS (High vs low) – FHS  Gao (2023)  maOR 0.69 (0.62-0.77, p<0.05) |  |
|  |  |  | AHEI (High vs low) – NHANES  Gao (2023)  maOR 0.80 (0.68- 0.93, p<0.05) |  |
|  |  |  | MDS (High vs low) – NHANES  Gao (2023)  maOR 0.76 (0.66, 0.87, p<0.05) |  |
|  | MASH prevalence |  | DASH (High vs low) – FHS  Gao (2023)  maOR 0.84 (0.72-0.98, p=0.02) | MDS (High vs low) – FHS  Gao (2023)  maOR 0.86 (0.74-1.01, p=0.06) |
|  |  |  | DASH (High vs low) – NHANES  Gao (2023)  maOR 0.73 (0.62-0.86, p=0.04) | MDS (High vs low) – NHANES  Gao (2023)  maOR 0.75 (0.6-0.95, p=0.09) |
|  |  |  | AHEI (High vs low) – FHS  Gao (2023)  maOR 0.84 (0.73-0.98, p=0.03) | AHEI (High vs low) – NHANES  Gao (2023)  maOR 0.78 (0.62-0.97, p=0.12) |
|  | Advanced Cirrhosis and Fibrosis prevalence | (poor vs excellent)  Unalp-Arida A (2023)  maOR 1.43 (1.00-2.05, p=0.04) | AHIE (Q5 vs Q1)  Park SY (2020)  maOR 0.64 (0.46-0.89, p=0.03) | AMD (Q5 vs Q1)  Park SY (2020)  maOR 1.02 (0.72-1.42, p=0.94) |
|  |  |  | DASH (Q5 vs Q1)  Park SY (2020)  maOR 0.68 (0.49-0.94, p=0.005) |  |
|  |  |  | HIE (Q5 vs Q1)  Park SY (2020)  maOR 0.74 (0.54-1.03, p=0.008) |  |
|  |  |  | DASH (High vs low) – FHS  Gao (2023)  maOR 0.77 (0.63-0.94, p=0.01) | DASH (High vs low) – NHANES  Gao (2023)  maOR 0.77 (0.63-0.94, p=0.08) |
|  |  |  | AHEI (High vs low) – FHS  Gao (2023)  maOR 0.81 (0.66-0.98, p=0.03) | AHEI (High vs low) – NHANES  Gao (2023)  maOR 0.83 (0.63-1.08, p=0.26) |
|  |  |  | MDS (High vs low) – FHS  Gao (2023)  maOR 0.78 (0.64-0.95, p=0.016) |  |
|  |  |  | MDS (High vs low) – NHANES  Gao (2023)  maOR 0.69 (0.58-0.82, p=0.02) |  |
|  | All-cause mortality |  | Yoo ER (2020) (Entire cohort)  maOR 0.81 (0.71-0.92, p=0.002) | Yoo ER (2020) (MASLD cohort)  maOR 0.98 (0.77-1.10, p=0.484) |
|  |  |  | Yoo ER (2020) (Non-NAFLD cohort)  maOR 0.91 (0.86-0.96, p=0.002) |  |
|  | Cancer-related mortality |  | Yoo ER (2020) (Entire cohort)  maOR 0.64 (0.48-0.85, p=0.003) | Yoo ER (2020) (Non-MASLD cohort)  maOR 0.88 (0.54-1.42, p=0.58) |
|  |  |  | Yoo ER (2020) (NAFLD cohort)  maOR 0.52 (0.35-0.80, p=0.004) |  |
|  | Cardiovascular disease |  |  | Yoo ER (2020) (Entire cohort)  maOR 0.90 (0.64-1.28, p=0.55) |
|  |  |  |  | Yoo ER (2020) (MASLD cohort)  maOR 0.97 (0.54-1.72, p=0.90) |
|  |  |  |  | Yoo ER (2020) (Non-MASLD cohort)  maOR 0.86 (0.60-1.24, p=0.41) |

| **SDOH** | **Outcome** | **Studies predicting increased risk** | **Studies predicting decreased risk** | **Studies predicting similar risk** |
| --- | --- | --- | --- | --- |
| **Food insecurity** | MASLD prevalence | (low vs high)  Golovaty (2020)  maOR 1.36 (1.02, 1.83, p<0.05) |  |  |
|  |  | (very low vs high)  Golovaty (2020)  maOR 1.52 (1.05, 2.18, p<0.05) |  |  |
|  | Advanced Cirrhosis and Fibrosis prevalence | (very low vs high)  Golovaty (2020)  maOR 3.64 (1.53, 8.65, p<0.01) |  | (low vs high)  Golovaty (2020)  maOR 1.39 (0.78, 2.51, p>0.05) |
|  | All-cause mortality | Kardashian A (2022) (NAFLD Cohort)  aHR 1.46 (1.08-1.97, p=0.01) |  |  |
|  |  | Kardashian A (2022) (AF Cohort)  aHR 1.37 (1.01-1.86, p=0.04) |  |  |
|  | Inpatient healthcare utilization |  |  | Kardashian A (2022) (MASLD Cohort)  aHR 1.46 (0.91-2.34, p=0.12) |
|  |  |  |  | Kardashian A (2022) (AF Cohort)  aHR 1.53 (0.83-2.81, p=0.17) |
|  | Outpatient healthcare utilization | Kardashian A (2022) (MASLD Cohort)  aHR 1.31 (1.05-1.63, p=0.02) |  | Kardashian A (2022) (AF Cohort)  aHR 1.09 (0.72-1.66, p=0.68) |

| **SDOH** | **Outcome** | **Studies predicting increased risk** | **Studies predicting decreased risk** | **Studies predicting similar risk** |
| --- | --- | --- | --- | --- |
| **Employment (vs full time)** | MASLD prevalence |  |  | Unemployment alone  Giammarino (2022)  uaOR 0.98 (0.85‐1.12, p=0.80) |
|  | MASH prevalence |  |  | Unemployment alone  Giammarino (2022)  uaOR 0.98 (0.80‐1.20, p=0.8) |
|  | Advanced Cirrhosis and Fibrosis prevalence |  |  | Unemployment alone  Giammarino (2022)  uaOR 0.84 (0.70‐1.02, p=0.079) |
|  | Fibrosis progression | Part time  Ruiz-Casas L (2021)  maOR 1.80 (1.02–3.13, p=0.03) |  | Self-employed  Ruiz-Casas L (2021)  maOR 0.91 (0.40–1.94, p=0.82) |
|  |  | Retired  Ruiz-Casas L (2021)  maOR 2.22 (1.07–4.51, p =0.03) |  | Student  Ruiz-Casas L (2021)  maOR 1.93 (0.39–7.08, p =0.36) |
|  |  | Unemployed  Ruiz-Casas L (2021)  maOR 2.32 (1.12–4.66, p =0.02) |  | Home-maker  Ruiz-Casas L (2021)  maOR 1.96 (0.73–4.79, p = 0.15) |

| **SDOH** | **Outcome** | **Studies predicting increased risk** | **Studies predicting decreased risk** | **Studies predicting similar risk** |
| --- | --- | --- | --- | --- |
| **Foreign born** | MASLD prevalence | Giammarino (2022)  uaOR 1.01 (1.00‐1.02, p=0.008) | Le (2020)  maOR 0.48 (0.33-0.70, p<0.001) |  |
|  | MASH prevalence |  |  | Giammarino (2022)  uaOR 1.00 (0.98‐1.01, p=0.74) |
|  | Advanced Cirrhosis and Fibrosis prevalence | Giammarino (2022)  uaOR 1.02 (1.01‐1.03, p=0.0003) |  |  |
|  | Received healthcare 1-3 times in past year |  |  | Le (2020)  maOR 1.03 (0.60-1.78, p=0.91) |
|  | Received healthcare 4-9 times in past year |  | Le (2020)  maOR 0.56 (0.34-0.93, p=0.02) |  |
|  | Received healthcare +10 times in past year |  | Le (2020)  maOR 0.41 (0.21-0.83, p=0.01) |  |

| **SDOH** | **Outcome** | **Studies predicting increased risk** | **Studies predicting decreased risk** | **Studies predicting similar risk** |
| --- | --- | --- | --- | --- |
| **Sedentary lifestyle/ Physical Inactivity** | MASLD prevalence | Unalp-Arida A (2023)  maOR 2.29 (1.30–4.03, p=0.006) | Chen W (2023) (Active vs inactive)  maOR 0.56 (0.46–0.68, p<0.001) | Unalp-Arida A (2023) (Inactive vs active)  maOR 1.26 (0.99–1.59, p=0.056) |
|  | Advanced Cirrhosis and Fibrosis prevalence | Unalp-Arida A (2023)  maOR 1.68 (1.10-2.56, p=0.01) |  |  |
|  | Mortality |  | Chen W (2023) (Active vs inactive)  maOR 0.77 (0.60–0.99, p=0.04 |  |

| **SDOH** | **Outcome** | **Studies predicting increased risk** | **Studies predicting decreased risk** | **Studies predicting similar risk** |
| --- | --- | --- | --- | --- |
| **Marital status** | MASLD prevalence |  |  | Single parent  Giammarino (2022)  uaOR 1.00 (0.99‐1.00, p=0.57) |
|  | MASH prevalence |  |  | Single parent  Giammarino (2022)  uaOR 0.99 (0.98‐1.00, p=0.38) |
|  | Advanced Cirrhosis and Fibrosis prevalence |  |  | Single parent  Giammarino (2022)  uaOR 1.00 (0.99‐1.00, p=0.76) |
|  |  |  |  | Never married  Rieman-Klinger (2024)  maOR 1.45 (0.32,6.47, p=0.62) |

| **SDOH** | **Outcome** | **Studies predicting increased risk** | **Studies predicting decreased risk** | **Studies predicting similar risk** |
| --- | --- | --- | --- | --- |
| **Affluence vs Disadvantage**  **Cohorts** | All cause mortality | Chen VL (2023) (Disadvantage cohort)  aHR 2.08 (1.54–2.81, p < 0.0001) | Chen VL (2023) (Affluence cohort)  aHR 0.49 (0.37-0.66, p < 0.0001) |  |
|  | Cardiovascular disease | Chen VL (2023) (Disadvantage cohort)  asHR 1.36 (1.10–1.68, p = 0.004) | Chen VL (2023) (Affluence cohort)  asHR 0.71 (0.57–0.88), p = 0.001) |  |
|  | Liver related events |  | Chen VL (2023) (Affluence cohort)  asHR 0.60 (0.39-0.92, p = 0.02) | Chen VL (2023) (Disadvantage cohort)  asHR 1.36 (0.90–2.07, p = 0.15) |

| **SDOH** | **Outcome** | **Studies predicting increased risk** | **Studies predicting decreased risk** | **Studies predicting similar risk** |
| --- | --- | --- | --- | --- |
| **High needs**  (Age >65 or <5 years, and women) | MASLD prevalence |  |  | Giammarino (2022)  uaOR 1.003 (0.997‐1.009, p=0.34) |
|  | MASH prevalence |  |  | Giammarino (2022)  uaOR 1.007 (0.998‐1.016, p=0.11) |
|  | Advanced Cirrhosis and Fibrosis prevalence | Giammarino (2022)  uaOR 1.011 (1.004‐1.019, p=0.003) |  |  |

| **SDOH** | **Outcome** | **Studies predicting increased risk** | **Studies predicting decreased risk** | **Studies predicting similar risk** |
| --- | --- | --- | --- | --- |
| **Transportation difficulty** | MASLD prevalence |  |  | Giammarino (2022)  uaOR 1.005 (1.000‐1.011, p=0.06) |
|  | MASH prevalence |  |  | Giammarino (2022)  uaOR 1.004 (0.995‐1.012, p=0.37) |

| **SDOH** | **Outcome** | **Studies predicting increased risk** | **Studies predicting decreased risk** | **Studies predicting similar risk** |
| --- | --- | --- | --- | --- |
| **Hospital region (vs north east)** | All-cause mortality | South  Adejumo AC (2019) (IH)  maOR 1.22 (1.01-1.48, p=0.02) |  | Midwest  Adejumo AC (2019) (IH)  maOR 1.18 (0.96-1.45, p=0.22) |
|  |  | West  Adejumo AC (2019) (IH)  maOR 1.41 (1.15-1.73, p<0.0001) |  |  |
|  | Unfavorable discharge disposition |  | South  Adejumo AC (2019)  maOR 0.80 (0.74-0.87, p<0.0001) | Midwest  Adejumo AC (2019)  maOR 0.99 (0.91-1.09, p>0.99) |
|  |  |  | West  Adejumo AC (2019)  maOR 0.90 (0.83-0.98, p=0.0102) |  |
|  | Hospital charges | West  Adejumo AC (2019)  maOR 1.42 (1.29-1.56, p<0.0001) | Midwest  Adejumo AC (2019)  maOR 0.90 (0.81-0.99, p=0.0219) | South  Adejumo AC (2019)  maOR 1.00 (0.91-1.10, p=1.00) |
|  | Increased length of stay |  | Midwest  Adejumo AC (2019)  maOR 0.95 (0.91-1.00, p=0.0385) | South  Adejumo AC (2019)  maOR 0.99 (0.95-1.03, p>0.99) |
|  |  |  | West  Adejumo AC (2019)  maOR 0.93 (0.89-0.98, p=0.0009) |  |

| **SDOH** | **Outcome** | **Studies predicting increased risk** | **Studies predicting decreased risk** | **Studies predicting similar risk** |
| --- | --- | --- | --- | --- |
| **Hospital teaching status (vs rural)** | All-cause mortality | Urban teaching  Adejumo AC (2019) (IH)  maOR 1.42 (1.14-1.76, p=0.0003) |  | Urban non-teaching  Adejumo AC (2019) (IH)  maOR 1.16 (0.93-1.44, p=0.33) |
|  | Unfavorable discharge disposition |  | Urban non-teaching  Adejumo AC (2019)  maOR 0.90 (0.93-0.97, p=0.003) |  |
|  |  |  | Urban teaching  Adejumo AC (2019)  maOR 0.83 (0.77-0.90, p<0.0001) |  |
|  | Hospital charges | Urban non-teaching  Adejumo AC (2019)  maOR 1.59 (1.49-1.70, p<0.0001) |  |  |
|  |  | Urban teaching  Adejumo AC (2019)  maOR 1.83 (1.70-1.96, p<0.0001) |  |  |
|  | Increased length of stay | Urban non-teaching  Adejumo AC (2019)  maOR 1.13 (1.09-1.17, p<0.0001) |  |  |
|  |  | Urban teaching  Adejumo AC (2019)  maOR 1.23 (1.19-1.27, p<0.0001) |  |  |

SDOH: social determinant of health; MASLD: metabolic dysfunction-associated steatotic liver disease; MASH: metabolic dysfunction-associated steatohepatitis; OR: odds ratio; NHANES: National Health and Nutrition Examination Survey; HS: high school; aaOR: Age-adjusted odds ratio; maOR: Multivariate-adjusted odds ratio; NH: Non-Hispanic; P:I : Poverty to income ratio; NH: non-Hispanic; LRE: liver related event; PE: pulmonary embolism; CVD: cardiovascular disease; AF: advanced fibrosis.

**Supplement** **Table 3: Quality Assessment of all included studies**

| Author & Year | 1 | 2 | 3 | 4 | 5 | 6 | 7 | 8 | 9 | 10 | 11 | 12 | 13 | 14 | Total |
| --- | --- | --- | --- | --- | --- | --- | --- | --- | --- | --- | --- | --- | --- | --- | --- |
| Chen VL, 2023 | Yes | Yes | NA | Yes | No | Yes | Yes | No | Yes | No | Yes | No | NA | Yes | 8 |
| Unalp-Arida A, 2022 | Yes | Yes | Yes | Yes | No | No | No | Yes | Yes | No | Yes | NA | NA | Yes | 8 |
| Kardashian A, 2022 | Yes | Yes | Yes | Yes | No | No | No | Yes | Yes | No | Yes | NA | NA | Yes | 8 |
| Giammarino, 2022 | Yes | Yes | No | Yes | No | Yes | Yes | Yes | Yes | No | Yes | NA | NA | Yes | 9 |
| Ruiz-Casas L, 2021 | Yes | Yes | Yes | Yes | No | No | No | Yes | Yes | No | Yes | NA | NA | Yes | 8 |
| Yoo ER, 2022 | Yes | Yes | NA | Yes | No | No | No | No | Yes | No | Yes | NA | NA | Yes | 6 |
| Golovaty I, 2020 | Yes | Yes | NA | Yes | No | No | No | Yes | Yes | No | Yes | NA | NA | Yes | 7 |
| Le MH, 2020 | Yes | Yes | NA | Yes | No | No | No | Yes | Yes | No | Yes | NA | NA | Yes | 7 |
| Park SY, 2020 | Yes | Yes | No | Yes | No | No | No | Yes | Yes | No | Yes | NA | NA | Yes | 7 |
| Adejumo AC, 2019 | Yes | Yes | Yes | Yes | No | No | No | Yes | Yes | No | Yes | NA | NA | Yes | 8 |
| Bambha K, 2012 | Yes | Yes | NA | Yes | No | No | No | No | Yes | No | Yes | Yes | NA | Yes | 7 |
| Alvi AT, 2023 | Yes | Yes | Yes | Yes | No | No | Yes | Yes | Yes | No | Yes | NA | NA | Yes | 9 |
| Gao V, 2023 | Yes | Yes | Yes | Yes | No | No | No | No | Yes | No | Yes | NA | NA | Yes | 7 |
| Chen W, 2023 | Yes | Yes | Yes | Yes | No | No | No | Yes | Yes | No | Yes | NA | NA | Yes | 8 |
| Hu E, 2022 | Yes | Yes | NA | Yes | No | No | No | Yes | Yes | No | Yes | NA | NA | Yes | 7 |
| Han MAT, 2021 | Yes | Yes | NA | Yes | No | No | No | No | Yes | No | Yes | NA | NA | NA | 5 |
| Rastegari S, 2019 | No | Yes | No | Yes | No | No | No | No | Yes | No | Yes | NA | NA | NA | 4 |
| Rieman-Klinger, 2024 | Yes | Yes | NA | Yes | No | No | No | Yes | Yes | No | Yes | NA | BA | Yes | 7 |

**Supplemental Figure 1: Funnel Plots to Assess for Publication Bias**

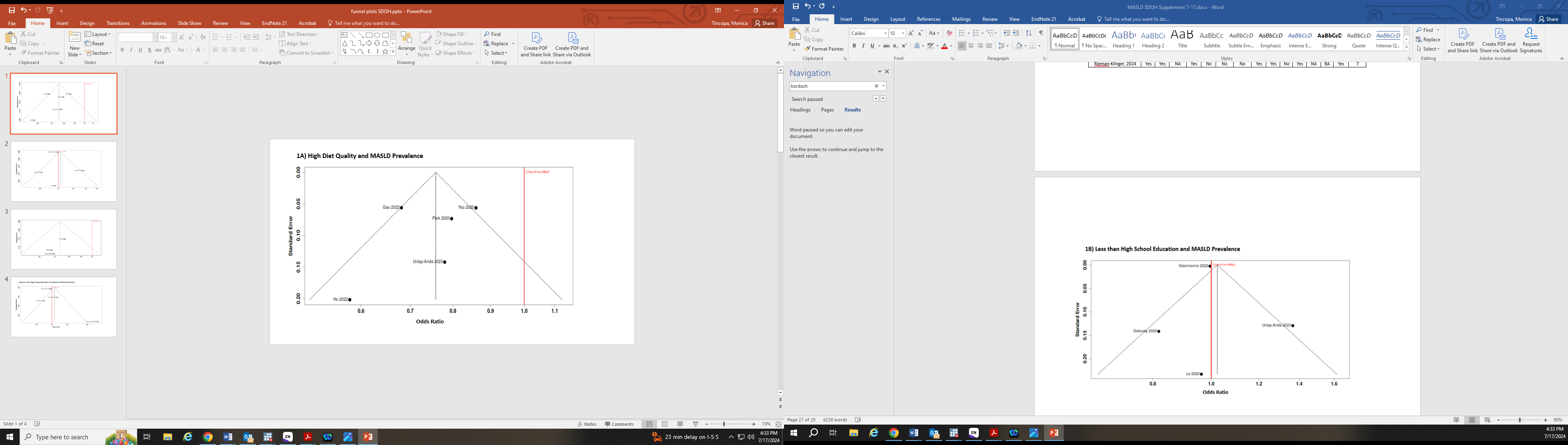

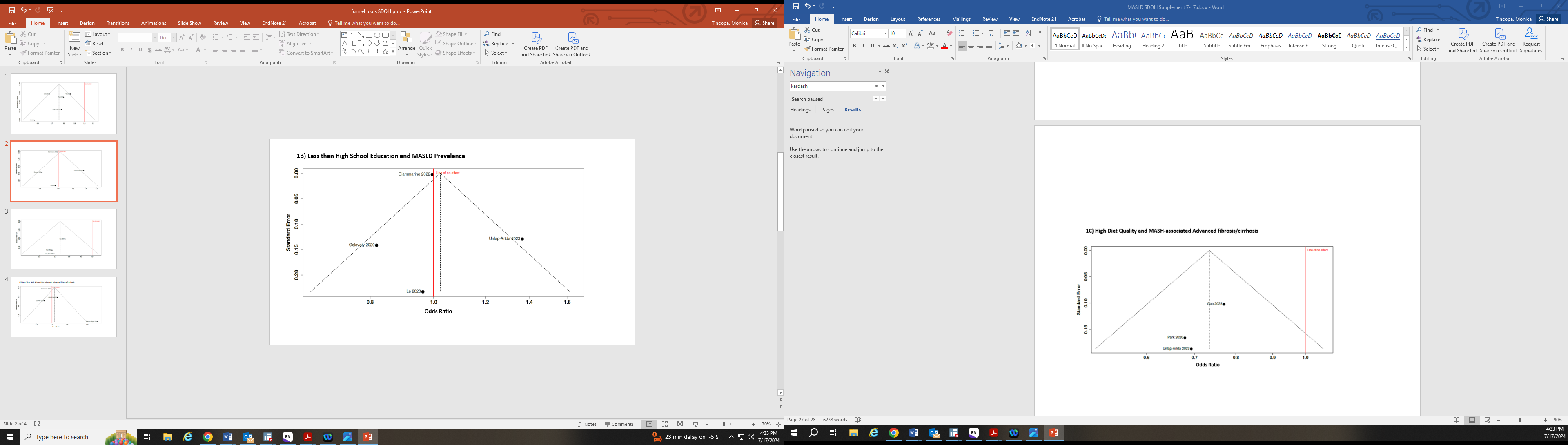

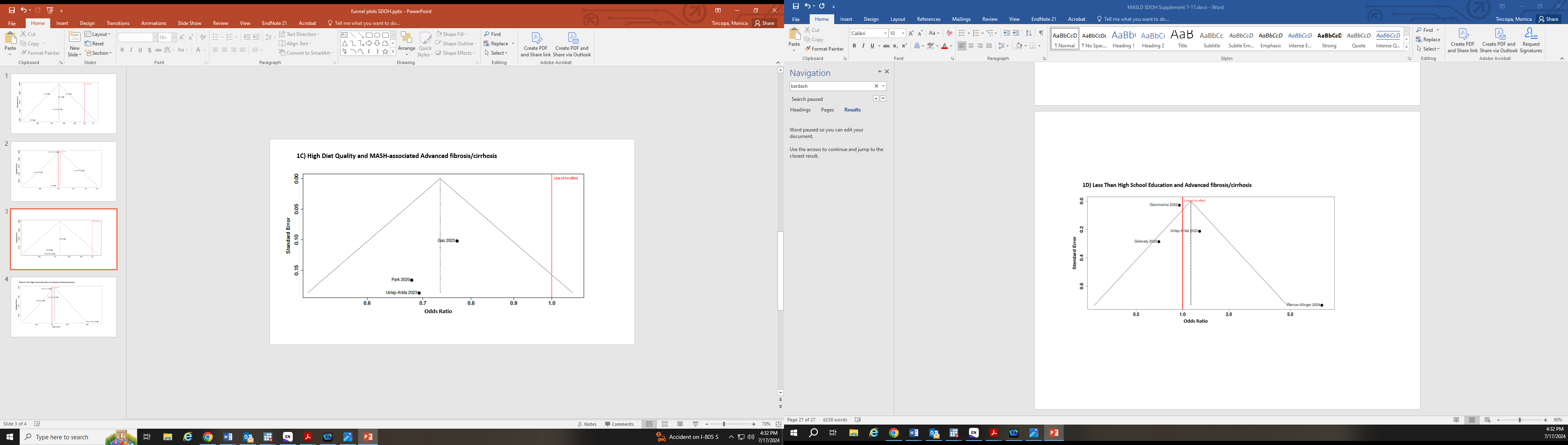

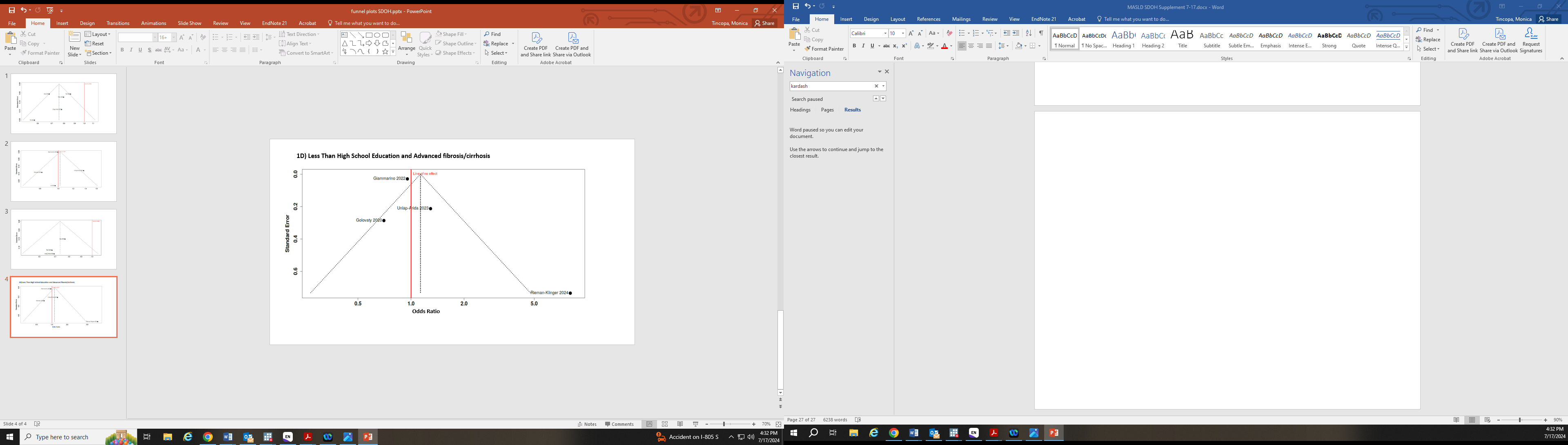
