## Supplementary material for "Social Determinants of Health Factors Associated with Metabolic Dysfunction-Associated Steatotic Liver Disease Prevalence and Severity: A Systematic Review and Meta-analysis": Tables

**Table 1: Studies Evaluating SDOH Factors and Association with MASLD Prevalence**

| **Author**  **(Year)** | **Study Design** | **Study**  **Cohort N=** | **Diagnostic Methods** | **SDOH Variables** | **OR (95% CI)** |
| --- | --- | --- | --- | --- | --- |
| Unalp-Arida A (2023) | Cross-sectional | NHANES 2017-2020, n=7923 | Fibroscan CAP (>300 dB/m) | 1- NH Black vs White race  2- NH Asian vs White race  3- Hispanic vs White race  4- Diet quality (poor vs excellent)  5- Physical inactivity  6- Sedentary behavior (>12hrs/day)  7- Education (Below HS vs college)  8- Education (HS vs college)  9- Income (P:I <1.65 vs >5)  10- Income (P:I 1.65-3.2 vs >5)  11- Smoking (Former vs never)  12- Smoking (Current vs never)  13- Health Insurance (Public vs Private)  14- Health Insurance (Uninsured vs Private) | 1- maOR 0.51 (0.39–0.65, **p<0.001)**  2- maOR 1.49 (1.05–2.10, **p=0.02**)  3- maOR 1.17 (0.86–1.59, p=0.31)  4- maOR 1.28 (0.97–1.69, p=0.07)  5- maOR 1.26 (0.99–1.59, p=0.06)  6- maOR 2.29 (1.30–4.03, **p=0.006)**  7- aaOR 1.36 (1.06-1.76, **p=0.02**)  8- aaOR 1.58 (1.24,2.01**, p<0.001**)  9- aaOR 1.36 (1.02-1.82, **p=0.03**)  10- aaOR 1.32 (0.94-1.85, p=0.11)  11- aaOR 1.40 (1.19–1.66, **p<0.001**)  12- aaOR 1.05 (0.83–1.33, p=0.65)  13- aaOR 1.08 (0.89-1.32, p=0.40)  14- aaOR 0.97 (0.78-1.21, p=0.78) |
| Giammarino  (2022) | Retrospective cohort | 12 hospitals in Northwell Health system, New York, n=614 | Liver biopsy | 1- Unemployment  2- Only high school diploma  3- Bachelor’s degree or higher  4- Private health care  5- Public health care  6- No health care  7- Foreign born  8- SDI  9- Poverty  10- Single parent  11- Black non‐Hispanic  12- High school dropout  13- No car  14- Nonemployment  15- High needs  16- Hispanic  17- Number of SDIs (1)  18- Number of SDIs (2)  19- Number of SDIs (3)  20- Number of SDIs (4)  21- Number of SDIs (5)  22- Number of SDIs (6) | 1- OR 0.98 (0.85‐1.12, p=0.80)  2- OR 0.99 (0.97‐1.01, p=0.69)  3- OR 1.00 (0.99‐1.01, p=0.93)  4- OR 0.99 (0.97‐1.00, p=0.14)  5- OR 1.00(0.98‐1.02, p=0.50)  6- OR 1.03(0.99‐1.07, p=0.14)  7- OR 1.01 (1.00‐1.02, **p=0.008**)  8- OR 1.00 (0.99‐1.00, p=0.14)  9- OR 1.00 (0.99‐1.01, p=0.17)  10- OR 1.00 (0.99‐1.00, p=0.57)  11- OR 1.00 (0.99‐1.00, p=0.97)  12- OR 1.00 (0.99‐1.00, p=0.19)  13- OR 1.00 (1.00‐1.01, p=0.06)  14- OR 1.00 (0.99‐1.00, p=0.96)  15- OR 1.00 (0.99‐1.00, p=0.34)  16- OR 1.00 (0.99‐1.01, p=0.20)  18- maOR 1.09 (0.61‐1.92, p=0.76)  19- maOR 1.24 (0.83‐1.84, p=0.27)  20- maOR 1.37 (0.94‐2.00, p=0.10)  21- maOR 1.39 (0.96‐2.03, p=0.07)  22- maOR 1.34 (0.92‐1.95, p=0.12)  23 - maOR 1.49 (1.03‐2.17, **p=0.03)** |
| Yoo  (2020) | Cross-sectional | NHANES (1988-1994), n=10858 | Ultrasound | 1- Diet quality (high vs poor) | 1- maOR 0.86 (0.77-0.96, **p=0.006**) |
| Golovaty  (2020) | Cross-sectional | NHANES (2005-2014), n=2627 | USFLI | 1- Food insecure (low vs high)  2- Food insecure (very low vs high)  3- Race (African American vs white)  4- Race (Mexican vs white)  5- Race (Non-Mexican Hispanic vs white)  6- Race (Other vs white)  7- Household income (<10,000 vs ≥25,000)  8- Household income (10,000–19,999 vs ≥25,000)  9- Household income (20,000–24,999 vs ≥25,000)  10- Education (<High school vs >high school)  11- Education (High school vs >high school)  12- Smoking (yes vs no) | 1- maOR 1.36 (1.02, 1.83**, p<0.05)**  2- maOR 1.52 (1.05, 2.18**, p<0.05)**  3- maOR 0.42 (0.31, 0.58**, p<0.001**)  4- maOR 2.03 (1.50, 2.74, **p<0.001**)  5- maOR 0.93 (0.68, 1.27, p>0.05)  6- maOR 0.50 (0.32, 0.77**, p<0.01**)  7- maOR 1.08 (0.76, 1.55, p>0.05)  8- maOR 0.99 (0.76, 1.29, p>0.05)  9- maOR 0.87 (0.62, 1.20, p>0.05)  10- maOR 0.82 (0.62, 1.08, p>0.05)  11- maOR 0.89 (0.67, 1.18, p>0.05)  12- maOR 1.04 (0.84, 1.28, p>0.05) |
| Le  (2020) | Cross-sectional | NHANES (2011-2016), n=4538 | USFLI | 1- Race (NH Black vs NH White)  2- Race (Mexican American vs NH White)  3- Race (Hispanic vs NH White)  4- Race (NH Asian vs NH White)  5- Race (Other vs NH White)  6- Foreign born (yes vs no)  7- Income to poverty ratio  8- Educational level (HS vs no HS)  9- Educational level (College vs no HS)  10- Smoking (Current/Ex vs Never) | 1- maOR 0.44 (0.31-0.62, **p<0.001**)  2- maOR 3.09 (2.07-4.60**, p<0.001**)  3- maOR 1.41 (0.79-2.50, p=0.23)  4- maOR 0.95 (0.51-1.77, p=0.87)  5- maOR 1.38 (0.65-2.94, p=0.39)  6- maOR 0.48 (0.33-0.70**, p<0.001**)  7- maOR 0.91 (0.82-1.00, p=0.06)  8- maOR 0.96 (0.61-1.52, p=0.86)  9- maOR 0.80 (0.48-1.36, p=0.40)  10- maOR 1.09 (0.78-1.51, p=0.60) |
| Park  (2020) | Case-control | Medicare claims (1999-2016), n=32251 | ICD codes | 1- Alternative Healthy Eating Index (Q5 vs Q1)  2- Alternate Mediterranean diet (Q5 vs Q1)  3- DASH (Q5 vs Q1)  4- Healthy Eating Index (Q5 vs Q1) | 1- maOR 0.96 (0.84-1.10, p=0.82)  2- maOR 1.00 (0.86-1.15, p=0.63)  3- maOR 0.80 (0.69-0.92, **p=0.005**)  4- maOR 0.84 (0.74-0.97, **p=0.03**) |
| Chen  (2023) | Retrospective cohort | NHANES 1988-1994 (n=9068) and NHANES 1999-2014 (n=14963) | USFLI | Physical activity (Active vs inactive) | 1- maOR 0.56 (0.46–0.68, **p<0.001**) |
| Hu  (2022) | Cross sectional | NHANES 2017-2018 (n=3486) | Fibroscan | Fast food per week (7 or more times vs none) | maOR 1.70 (1.15-2.51**, p=0.01**) |
| Gao  (2023) | Cross sectional | NHANES 2017-2018 (n=3295) and Framingham Heart Study (FHS) (n=2532) | Fibroscan | Diet quality (High vs low) – FHS  1- DASH  2- AHEI  3- MDS  Diet quality (High vs low) – NHANES  1- DASH  2- AHEI  3- MDS | 1- maOR 0.68 (0.61-0.76, **p<0.05)**  2- maOR 0.66 (0.59-0.73, **p<0.05)**  3- maOR 0.69 (0.62-0.77, **p<0.05)**  1- maOR 0.84 (0.71-1.00, p>0.05)  2- maOR 0.80 (0.68- 0.93, **p<0.05)**  3- maOR 0.76 (0.66, 0.87, **p<0.05)** |

SDOH: social determinant of health; MASLD: metabolic dysfunction-associated steatotic liver disease; OR: odds ratio; NHANES: National Health and Nutrition Examination Survey; CAP: controlled attenuation parameter; HS: high school; aaOR: Age-adjusted odds ratio; maOR: Multivariate-adjusted odds ratio; NH: Non-Hispanic; P:I : Poverty to income ratio; SDI: social deprivation index; USFLI: ultrasonographic fatty liver indicator; NH: non-Hispanic; DASH: Dietary Approaches to Stop Hypertension; AHEIL Alternative Healthy Eating Index; MDS: Mediterranean Diet Serving; FHS: Framingham Heart Study.

**Table 2: Studies Evaluating SDOH Factors and Association with MASH Prevalence**

| **Author**  **(Year)** | **Study Design** | **Study**  **Cohort**  **N=** | **Diagnostic Methods** | **SDOH Variables** | **OR (95% CI)** |
| --- | --- | --- | --- | --- | --- |
| Giammarino  (2022) | Retrospective cohort | 12 hospitals in Northwell Health system, New York, n=614 | Liver biopsy | 1- Unemployed  2- Only high school diploma  3- Bachelor’s degree or higher  4- Private health care  5- Public health care  6- No health care  7- Foreign born  8- SDI  9- Poverty  10- Single parent  11- Black non‐Hispanic  12- High school dropout  13- No car  14- Nonemployment  15- High needs  16- Hispanic  17- Foreign born  18- Number of SDIs (1)  19- Number of SDIs (2)  20- Number of SDIs (3)  21- Number of SDIs (4)  22- Number of SDIs (5)  23- Number of SDIs (6) | 1- OR 0.98 (0.80‐1.20, p=0.89)  2- OR 0.96 (0.94‐0.99, **p=0.03**)  3- OR 1.01 (1.00‐1.03**, p=0.02**)  4- OR 0.99 (0.98‐1.01, p=0.85)  5- OR 1.00 (0.97‐1.03, p=0.77)  6- OR 0.99 (0.93‐1.05, p=0.86)  7- OR 1.00 (0.98‐1.01, p=0.74)  8- OR 1.00 (0.93‐1.00, p=0.88)  9- OR 1.00 (0.99‐1.01, p=0.52)  10- OR 0.99 (0.98‐1.00, p=0.38)  11- OR 0.99 (0.98‐1.00, p=0.20)  12- OR 0.99 (0.98‐1.00, p=0.37)  13- OR 1.00 (0.99‐1.01, p=0.37)  14- OR 1.00 (0.98‐1.01, p=0.94)  15- OR 1.00 (0.99‐1.01, p=0.11)  16- OR 0.99 (0.98‐1.01, p=0.72)  17- OR 0.99 (0.98‐1.01, p=0.89)  18- maOR 1.24 (0.59‐2.61, p=0.55)  19- maOR 1.07 (0.65‐1.76, p=0.77)  20- maOR 1.6 (0.98‐2.61, p=0.05)  21- maOR 1.7 (1.09‐2.85, **p=0.01)**  22- maOR 1.94 (1.20‐3.14, **p=0.006)**  23- maOR 1.74 (1.09‐2.78, **p=0.01**) |
| Bambha  (2012) | Retrospective cohort | NASH CRN 2004 to 2008, n=1026 | Liver biopsy | 1- Race (Latino vs non-latino white)  2- Education level (>HS vs <HS)  3- Income (>50,000 vs <50,000)  4- Physical activity | 1- maOR 1.43 (0.88-2.33, p=0.15)  2- OR 0.74 (0.55-1.00, **p=0.05**)  3- OR 0.64 (0.49-0.85, **p<0.01)**  4- OR 1.00 (0.997-1.003, p=0.94) |
| Gao  (2023) | Cross sectional | NHANES 2017-2018 (n=3295) and Framingham Heart Study (FHS) (n=2532) | Fibroscan | Diet quality (High vs low) – FHS  1- DASH  2- AHEI  3- MDS  Diet quality (High vs low) – NHANES  1- DASH  2- AHEI  3- MDS | 1- maOR 0.84 (0.72-0.98, **p=0.02)**  2- maOR 0.84 (0.73-0.98**, p=0.03**)  3- maOR 0.86 (0.74-1.01, p=0.06)  1- maOR 0.73 (0.62-0.86, **p=0.04)**  1- maOR 0.78 (0.62-0.97, p=0.12)  1- maOR 0.75 (0.6-0.95, p=0.09) |

SDOH: social determinant of health; MASH: metabolic dysfunction-associated steatohepatitis; OR, odds ratio; NHANES: National Health and Nutrition Examination Survey; HS: high school; maOR: Multivariate-adjusted odds ratio; SDI: social deprivation index; DASH: Dietary Approaches to Stop Hypertension; AHEIL Alternative Healthy Eating Index; MDS: Mediterranean Diet Serving; FHS: Framingham Heart Study.

**Table 3: Studies Evaluating SDOH Factors and Association with MASH-Associated Advanced Fibrosis/Cirrhosis**

| **Author**  **(Year)** | **Study Design** | **Study**  **Cohort**  **N=** | **Diagnostic Methods** | **SDOH Variables** | **OR (95% CI)** |
| --- | --- | --- | --- | --- | --- |
| Unalp-Arida (2023) | Cross-sectional | NHANES 2017-2020, n=6776 | Fibroscan (>8 kPa) | 1- NH Black vs White race  2- NH Asian vs White race  3- Hispanic vs White race  4- Diet quality (poor vs excellent)  5- Physical inactivity  6- Sedentary behavior (>12hrs/day)  7- Education (Below HS vs college)  8- Education (HS vs college)  9- Income (P:I <1.65 vs >5)  10- Income (P:I 1.65-3.2 vs >5)  11- Smoking (Former vs never)  12- Smoking (Current vs never)  13- Health Insurance (Public vs Private)  14- Health Insurance (Uninsured vs Private) | 1- maOR 0.78 (0.53-1.15, p=0.20)  2- maOR 1.21 (0.77-1.90, p=0.40)  3- maOR 0.94 (0.65-1.35, p=0.72)  4- maOR 1.43 (1.00-2.05, **p=0.04**)  5- aaOR 1.45 (1.19-1.77**, p<0.001**)  6- maOR 1.68 (1.10-2.56**, p=0.01)**  7- maOR 1.29 (0.85-1.96, p=0.22)  8- maOR 1.77 (1.05-3.00, **p=0.03)**  9- maOR 2.00 (1.18-3.40, **p=0.01**)  10- maOR 2.59 (1.38-4.85**, p=0.005**)  11- aaOR 1.30 (1.03-1.64, **p=0.02**)  12- aaOR 1.12 (0.90-1.39, p=0.30)  13- aaOR 1.06 (0.81-1.39, p=0.66)  14- aaOR 1.04 (0.72-1.50, p=0.84) |
| Giammarino  2022 | Retrospective cohort | 12 hospitals in Northwell Health system, New York, n=614 | Liver biopsy | 1- Unemployed  2- Only high school diploma  3- Bachelor’s degree or higher  4- Private health care  5- Public health care  6- No health care  7- Foreign born  8- SDI  9- Poverty  10- Single parent  11- Black non‐Hispanic  12- High school dropout  13- No car  14- Nonemployment  15- High needs  16- Hispanic  17- Foreign born  18- Number of SDIs (1)  19- Number of SDIs (2)  20- Number of SDIs (3)  21- Number of SDIs (4)  22- Number of SDIs (5)  23- Number of SDIs (6) | 1- OR 0.84 (0.70‐1.02, p=0.07)  2- OR 0.99 (0.97‐1.02, p=0.69)  3- OR 1.00 (0.98‐1.01, p=0.83)  4- OR 0.98 (0.96‐0.99**, p=0.04**)  5- OR 1.02 (1.00‐1.05, **p=0.03**)  6- OR 1.03 (0.98‐1.08, p=0.17)  7- OR 1.02 (1.01‐1.03**, p=0.0003)**  8- OR 1.00 (1.00‐1.01, p=0.06)  9- OR 1.00 (0.99‐1.01, p=0.14)  10- OR 1.00 (0.99‐1.00, p=0.76)  11- OR 0.99 (0.99‐1.00, p=0.73)  12- OR 1.00 (0.99‐1.01, p=0.06)  13- OR 1.01 (1.00‐1.01, **p=0.001)**  14- OR 0.99 (0.98‐1.00, p=0.80)  15- OR 1.01 (1.00‐1.01, **p=0.003)**  16- OR 1.00 (0.99‐1.01, p=0.13)  17- OR 1.02 (1.01‐1.03**, p=<0.001)**  18- maOR 1.41 (0.66‐3.01, p=0.36)  19- maOR 1.10 (0.67‐1.81, p=0.69)  20- maOR 1.54 (0.95‐2.49, p=0.07)  21- maOR 1.42 (0.89‐2.26, p=0.14)  22- maOR 1.49 (0.93‐2.38, p=0.09)  23- maOR 1.49 (0.93‐2.37, p=0.09) |
| Golovaty  (2020) | Cross-sectional | NHANES (2005-2014), n=2627 | NFS | 1- Food insecure (low vs high) (n=27)  2- Food insecure (very low vs high) (n=17)  3- Race (African American vs white) (n=173)  4- Race (Mexican vs white)  5- Race (Non-Mexican Hispanic vs white)  6- Race (Other vs white)  7- Household income (<10,000 vs ≥25,000)  8- Household income (10,000–19,999 vs ≥25,000)  9- Household income (20,000–24,999 vs ≥25,000)  10- Education (<High school vs >high school)  11- Education (High school vs >high school)  12- Smoking (yes vs no) | 1- maOR 1.39 (0.78, 2.51, p>0.05)  2- maOR 3.64 (1.53, 8.65**, p<0.01)**  3- maOR 1.00 (0.60, 1.67, p>0.05)  4- maOR 1.46 (0.78, 2.70, p>0.05)  5- maOR 0.32 (0.16, 0.66, **p<0.05)**  6- maOR 0.44 (0.16, 1.18, p>0.05)  7- maOR 2.29 (0.99, 5.32, p>0.05)  8- maOR 1.58 (0.77, 3.22, p>0.05)  9- maOR 1.31 (0.66, 2.60, p>0.05)  10- maOR 0.79 (0.43, 1.44, p>0.05)  11- maOR 0.70 (0.40, 1.23, p>0.05)  12- maOR 0.74 (0.46, 1.17, p>0.05) |
| Le  (2020) | Cross-sectional | NHANES (2011-2016), n=4538 | NFS | 1- Race (NH Black vs NH White)  2- Race (Mexican American vs NH White)  3- Race (Hispanic vs NH White)  4- Race (NH Asian vs NH White)  5- Race (Other vs NH White)  6- Smoking (Current/Ex vs Never) | 1- maOR 1.43 (0.85-2.39, p=0.17)  2- maOR 0.40 (0.24-0.67**, p=0.001)**  3- maOR 1.03 (0.46-2.28, p=0.94)  4- maOR 0.12 (0.29-0.50**, p=0.005)**  5- maOR 0.64 (0.35-1.19, p=0.15)  6- maOR 1.30 (0.85-1.99, p=0.22) |
| Park  (2020) | Case-control | Medicare claims (1999-2016), n=32251 | ICD codes | 1- Alternative Healthy Eating Index (Q5 vs Q1)  2- Alternate Mediterranean diet (Q5 vs Q1)  3- DASH (Q5 vs Q1)  4- Healthy Eating Index (Q5 vs Q1) | 1- maOR 0.64 (0.0.46-0.89, **p=0.03)**  2- maOR 1.02 (0.72-1.42, p=0.94)  3- maOR 0.68 (0.49-0.94**, p=0.005)**  4- maOR 0.74 (0.54-1.03, **p=0.008)** |
| Bambha  (2012) | Retrospective cohort | NASH CRN 2004-2008, n=1026 | Liver Biopsy | 1- Race (Latino vs non-latino white) | 1- maOR 0.78 (0.42-1.45, p=0.43) |
| Gao  (2023) | Cross sectional | NHANES 2017-2018 (n=3295) and Framingham Heart Study (FHS) (n=2532) | Fibroscan | Diet quality (High vs low) – FHS  1- DASH  2- AHEI  3- MDS  Diet quality (High vs low) – NHANES  1- DASH  2- AHEI  3- MDS | 1- maOR 0.77 (0.63-0.94, **p=0.01)**  2- maOR 0.81 (0.66-0.98, **p=0.03)**  3- maOR 0.78 (0.64-0.95, **p=0.016)**  1- maOR 0.77 (0.63-0.94, p=0.08)  1- maOR 0.83 (0.63-1.08, p=0.26)  1- maOR 0.69 (0.58-0.82**, p=0.02)** |
| Rieman-Klinger (2024) | Cross sectional | USCD cohort (n=264) | MRE | 1- Education (No college degree)  2- Martial status (Never married)  3- Income (<30k vs >30k)  4- Ethnicity (Hispanic) | 1- maOR 8.04 (1.9–33.85, **p=0.005**)  2- maOR 1.45 (0.32,6.47, p=0.62)  3- maOR 0.26 (0.08–0.85, **p=0.027**)  4- maOR 1.79 (0.63–5.07, p=0.27) |

SDOH: social determinant of health; MASH: metabolic dysfunction-associated steatohepatitis; OR, odds ratio; NHANES: National Health and Nutrition Examination Survey; HS: high school; aaOR: Age-adjusted odds ratio. maOR: Multivariate-adjusted odds ratio. NH: Non-Hispanic. P:I : Poverty to income ratio; SDI: social deprivation index; MRE: MR elastography; NFS: NAFLD fibrosis score; NH: non-Hispanic; DASH: Dietary Approaches to Stop Hypertension; AHEIL Alternative Healthy Eating Index; MDS: Mediterranean Diet Serving; FHS: Framingham Heart Study; UCSD: University of California San Diego.
